# Cerebrospinal fluid hemolysis precedes secondary brain injury after aneurysmal subarachnoid hemorrhage

**DOI:** 10.64898/2026.09.08.26362434

**Authors:** Raphael M. Buzzi, Kevin Akeret, Sandra Wymann, Sandra Mena Pérez, Damian Brülhart, Moritz Saxenhofer, Daniel Couto, Jasmin Fischer, Andreas Wassmer, Pavel Tafo, Svenja Ewert, Sara Täschler, Marlies Illi, Jacqueline Adam, Adrien Schmid, Ulrike Held, Florence Vallelian, Michael Hugelshofer, Thomas Gentinetta, Dominik J. Schaer

## Abstract

**BACKGROUND:** After aneurysmal subarachnoid hemorrhage (aSAH), erythrocyte lysis within cerebrospinal fluid (CSF) releases cell-free hemoglobin, a potential driver of secondary brain injury (SBI). Whether hemolysis precedes SBI and how its temporal associations compare with those of inflammation and neuroglial injury remain unclear.

**METHODS:** In this biomarker profiling study of the prospective multicenter HeMoVal cohort, 259 patients with aSAH requiring external ventricular drainage contributed 2,411 serial CSF samples over days 1–14 after hemorrhage. We quantified 22 analytes using spectrophotometry, immunoassay, and mass spectrometry; 19 were assigned to five prespecified disease pathway composites. Day- and site-adjusted generalized estimating equations estimated associations with same-day SBI and future first SBI (angiographic vasospasm, delayed cerebral ischemia, or delayed ischemic neurological deficit); a day-wise landmark analysis assessed robustness.

**RESULTS:** On the day of sampling, SBI was associated with four pathways, with the largest estimates for inflammation and neuroglial injury. Within the prespecified +72-hour horizon, only hemolysis (OR 1.40 per SD; 95% CI, 1.17–1.68) and the erythrophagocyte response (OR 1.28; 1.07–1.54) had FDR-significant forward associations with first SBI. Hemolysis had the largest forward estimate at every horizon; landmark analysis was concordant (pooled OR 1.86; 95% bootstrap CI, 1.37–2.65). The protective scavenger proteins haptoglobin and hemopexin declined to fitted plateaus of 2.4% and 12.5% of day-1 concentrations, versus ∼36% for plasma-protein references.

**CONCLUSION:** In a scavenger-depleted CSF compartment, hemolysis and the erythrophagocyte response were associated with subsequent SBI. These findings identify hemolysis as a candidate upstream process and support testing sustained CSF hemoglobin scavenging to prevent SBI.

**TRIAL REGISTRATION:** ClinicalTrials.gov NCT04998370.

**FUNDING:** CSL Behring; Swiss National Science Foundation; University of Zurich

## INTRODUCTION

Secondary brain injury (SBI) is a major contributor to death and long-term disability in patients who survive the initial bleed after aneurysmal subarachnoid hemorrhage (aSAH) (1, 2). The SBI composite evaluated here comprises angiographic vasospasm, delayed cerebral ischemia, and delayed ischemic neurological deficit, complications that typically develop between days 4 and 14 after bleeding (3, 4). After aneurysm rupture, erythrocytes accumulate within the cerebrospinal fluid (CSF) compartment(5) and undergo delayed lysis over days, releasing cell-free hemoglobin directly into the environment in which SBI evolves(6). This makes aSAH an unusually informative disease model for compartmental hemolysis, as the CSF compartment can be sampled repeatedly in patients requiring external ventricular drainage (EVD), providing longitudinal access to the evolving biology of hemolysis and its downstream consequences(7, 8).

Cell-free hemoglobin is biologically well positioned to contribute to secondary injury after aSAH(9). Extracellular hemoglobin depletes nitric oxide, promotes oxidative stress, and amplifies inflammatory and tissue-injury responses(7, 10–14). Experimental and translational studies support a mechanistic role for cell-free hemoglobin in vasospasm, oxidative injury, neuroinflammation, and neuronal dysfunction, and show that the plasma scavenger proteins haptoglobin and hemopexin buffer hemoglobin-mediated toxicity(15–19). In ex vivo studies, cell-free hemoglobin in patient CSF impaired nitric oxide–mediated dilation and promoted cerebral arterial constriction; haptoglobin reversed these effects(16). Convergent human evidence from iron-deposition imaging, prior CSF cohorts, and genetic studies points in the same direction (20–23). The unresolved clinical question is whether CSF hemolysis precedes the first occurrence of SBI and how its temporal associations compare with those of inflammation and neuroglial injury, two key injury-related processes.

The prospective multicenter HeMoVal study provided a serial-CSF platform to address this question (24). The registered primary analysis, reported separately, did not support spectrophotometric CSF oxyhemoglobin as an individual bedside monitor for the protocol-defined SBI composite (25). The present secondary analysis addresses a different question: how CSF disease pathways relate to the subsequent onset of SBI. The protocol had prospectively mandated daily CSF biobanking with standardized processing to enable biomarker-based disease pathway analyses (24). Guided by a consensus pathobiological framework (26), we quantified 22 analytes in 2,411 serial CSF samples across spectrophotometric, immunoassay, and study-specific targeted mass spectrometry platforms. Nineteen analytes were assigned a priori to five pathway composites: hemolysis, erythrophagocyte response, inflammation, neuroglial injury, and plasma proteins. The erythrophagocyte-response composite captured macrophage-associated heme/iron handling, whereas the plasma-protein composite served as a comparator for plasma-entry and scavenger-protein kinetics (27–30).

Collectively, dense longitudinal sampling and a study-specific mass-spectrometry panel quantifying erythrocyte-lysis markers independently of hemoglobin’s oxidation state enabled direct comparison of five prespecified pathways within the same cohort and risk period. We contrasted same-day associations with forward first-event associations to distinguish pathways accompanying SBI from those associated with subsequent SBI in patients who were event-free at sampling.

## RESULTS

### Five CSF pathways follow distinct temporal courses over the first 14 days

SBI was assessed daily at the bedside according to the published protocol(24) in a prospective manner. The clinical database was locked before any biomarker was measured (**Figure 1**), and the biomarker panel itself was designed by a team not involved in running the clinical study.

**Figure 1.**
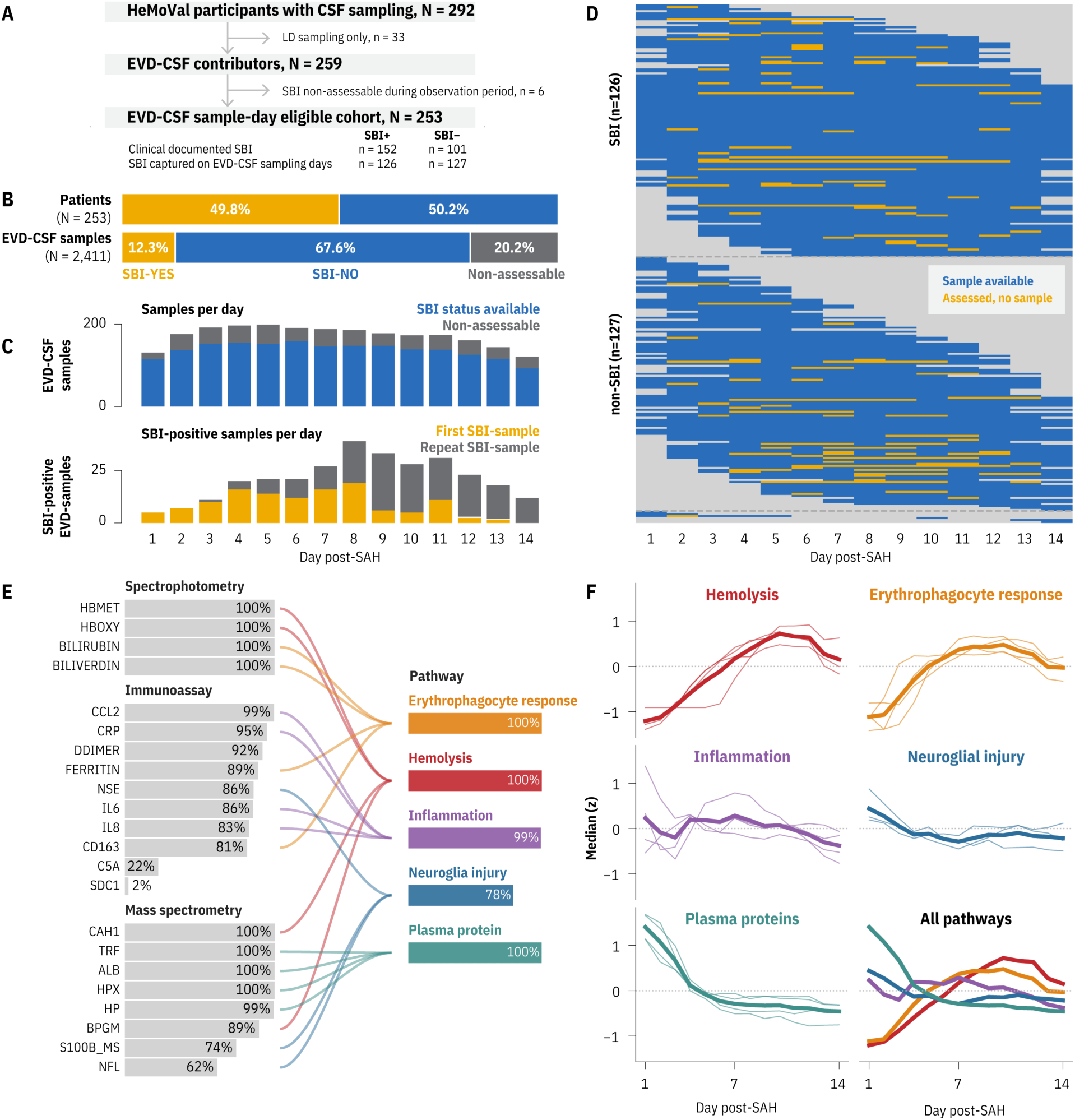
Participant composition, sampling structure, biomarker coverage, and longitudinal biomarker profiles of the HeMoVal EVD-CSF sample collection. **(A)** Of 292 HeMoVal patients providing any CSF sample (external ventricular or lumbar drain), 33 with lumbar-drain–only sampling were excluded, yielding the EVD cohort analyzed here. The EVD-sample cohort comprised 259 patients with aneurysmal subarachnoid hemorrhage (aSAH) across 8 sites (126 with SBI captured on an EVD-CSF sampling day, 127 with no SBI-positive sampling day, 6 with no EVD-CSF sample linked to an assessable same-day SBI status), contributing 2,411 valid CSF samples over days 1–14 after hemorrhage. Clinical SBI status, ascertained from the complete record, was documented in 152 of the 253 patients with assessable sample-day status and absent in 101; the 26-patient difference from the 126 above developed SBI only on days without EVD-CSF sampling. The inferential parent set comprised 1,925 patient-days from 253 patients with paired biomarker measurements and assessable SBI status; 486 non-assessable patient-days were retained for descriptive trajectory displays but withheld from inferential modeling. This parent set is used in full only by the same-day analysis (Figure 2); the first-event and landmark analyses draw horizon-specific subsets of it (see Methods). **(B)** Of the 253 patients in the inferential set, 126 (49.8%) had SBI captured on at least one EVD-CSF sampling day and 127 (50.2%) did not; across all 259 EVD patients, the 2,411 CSF samples are partitioned into SBI-positive sample-day (12.3%), assessable SBI-negative sample-day (67.6%), and non-assessable (20.2%) fractions. **(C)** CSF samples per day (assessable versus non-assessable) and SBI-positive EVD-CSF samples per day across days 1–14, split into each patient’s first SBI-positive EVD-CSF sample and any subsequent SBI-positive samples. This split indexes sampling, not clinical event onset: a patient’s first SBI-positive sample is the earliest sampled day on which SBI was recorded, which need not be the day the first clinical event occurred. **(D)** Patient-by-day sampling heatmap. Each row is a patient and each column a day after hemorrhage; cells indicate a sample available, no sample within that patient’s observed sampling span, or a day outside the observed sampling span. Patients are stratified by whether SBI was captured on at least one EVD-CSF sampling day (n = 126), on none (n = 127), or whether sample-day SBI status was non-assessable throughout (n = 6). These groupings classify sampling days, not clinical SBI status; clinical SBI was documented in 152 patients and absent in 101. **(E)** Biomarker-to-pathway map. Biomarkers are grouped by analytical platform and assigned to the five prespecified pathway composites. Bar length indicates the proportion of valid-day EVD samples with a measured and valid value. Ribbons link only the 19 analytes retained in the composites; C5a and syndecan-1, which fell below the 50% valid-measurement threshold, and D-dimer, which belongs to none of the five pathways, are shown with their coverage but carry no ribbon. The mass-spectrometry block denotes the study-specific MRM panel. **(F)** Analyte trajectories over days 1–14, grouped by prespecified pathway. Thin lines are the daily median z-score of each constituent analyte; the bold line is the unweighted mean of those constituent medians. The final panel overlays the five pathway-group summaries for direct comparison. These curves are not the pathway composite scores used in Figures 2–4, which take a row-wise mean per patient-day and are then re-standardized; the two are constructed differently and do not agree numerically.

The EVD-CSF biomarker analysis included 259 patients with aneurysmal subarachnoid hemorrhage who required external ventricular drainage across eight sites. Daily CSF biobanking yielded 2,411 EVD-derived CSF samples covering days 1–14 after hemorrhage. Among these, 1,925 patient-days from 253 patients had paired biomarker measurements and assessable SBI status and formed the parent set for inferential analyses, whereas 486 patient-days with non-assessable SBI status were retained for descriptive trajectory displays only (**Figure 1A–C**).

Per-sample biomarker availability was comparable across patients, although those with no SBI-positive sampling day contributed fewer sampling days (median 9 samples [IQR 5–11] for patients without an SBI-positive EVD-CSF sampling day vs 12 [IQR 9–14] for patients with one) due to faster EVD-weaning (**Figure 1D**). Among the 19 analytes retained in the pathway composites, per-analyte availability across the 2,411 sample-days ranged from 62% (NfL) to >99% (spectrophotometric hemolysis markers), and the ordering of availability tracked assay platform rather than outcome.

The biomarker panel comprised 22 analytes measured by spectrophotometry, immunoassays, and an analytically qualified 8-plex multiple-reaction monitoring mass spectrometry (MRM)-panel, developed specifically for this study. These biomarkers captured erythrocyte, neuroglial, and plasma proteins, including the scavengers haptoglobin and hemopexin, and were grouped into five biological pathways (**Figure 1E**). For inferential models, each pathway was represented by the current-day mean of its constituent biomarker z-scores, re-standardized within the relevant analysis set. Nineteen of the 22 analytes entered the five composites. C5a and syndecan-1 fell below the prespecified 50% valid-measurement threshold, and D-dimer, although measured reliably, belongs to none of the five pathways; all three are reported in the deposited dataset (Zenodo, https://doi.org/10.5281/zenodo.20733070).

Grouping the analyte trajectories by pathway revealed distinct temporal signatures over days 1– 14 (**Figure 1F**). Each panel shows the daily median z-score of every constituent analyte, summarized by their unweighted mean: the hemolysis and erythrophagocyte-response analytes rose late, the plasma proteins peaked early and declined, and the inflammation and neuroglial-injury analytes showed no monotonic trend. Within the hemolysis, erythrophagocyte-response and plasma-protein groups the constituent analytes moved together and closely tracked their group summary, whereas the inflammation and neuroglial-injury constituents were more heterogeneous.

### Four of five CSF pathways are associated with same-day SBI

We first examined which CSF pathways were associated with first or recurrent SBI on the same day as sample collection, using all clinically assessable patient-days (including days after a patient’s first event). Four of the five prespecified pathways were significantly associated with same-day SBI status after Benjamini–Hochberg correction across pathways, in a generalized estimating equations (GEE) model with exchangeable working correlation, robust sandwich standard errors, and day and site fixed effects (**Figure 2A**). Same-day associations were largest for inflammation (OR 1.47, 95% CI 1.26–1.73, q < 0.001) and neuroglial injury (OR 1.47, 95% CI 1.21–1.78, q < 0.001), followed by hemolysis (OR 1.31, 95% CI 1.06–1.63, q = 0.021) and the erythrophagocyte response (OR 1.25, 95% CI 1.02–1.54, q = 0.041), whereas the plasma-protein composite was not significantly associated with same-day SBI (**Figure 2A**). Model diagnostics did not suggest major pathway-specific misspecification (**Supplemental Figure 1**) and estimates were concordant under an AR (1) working-correlation sensitivity (**Supplemental Figure 2**).

**Figure 2.**
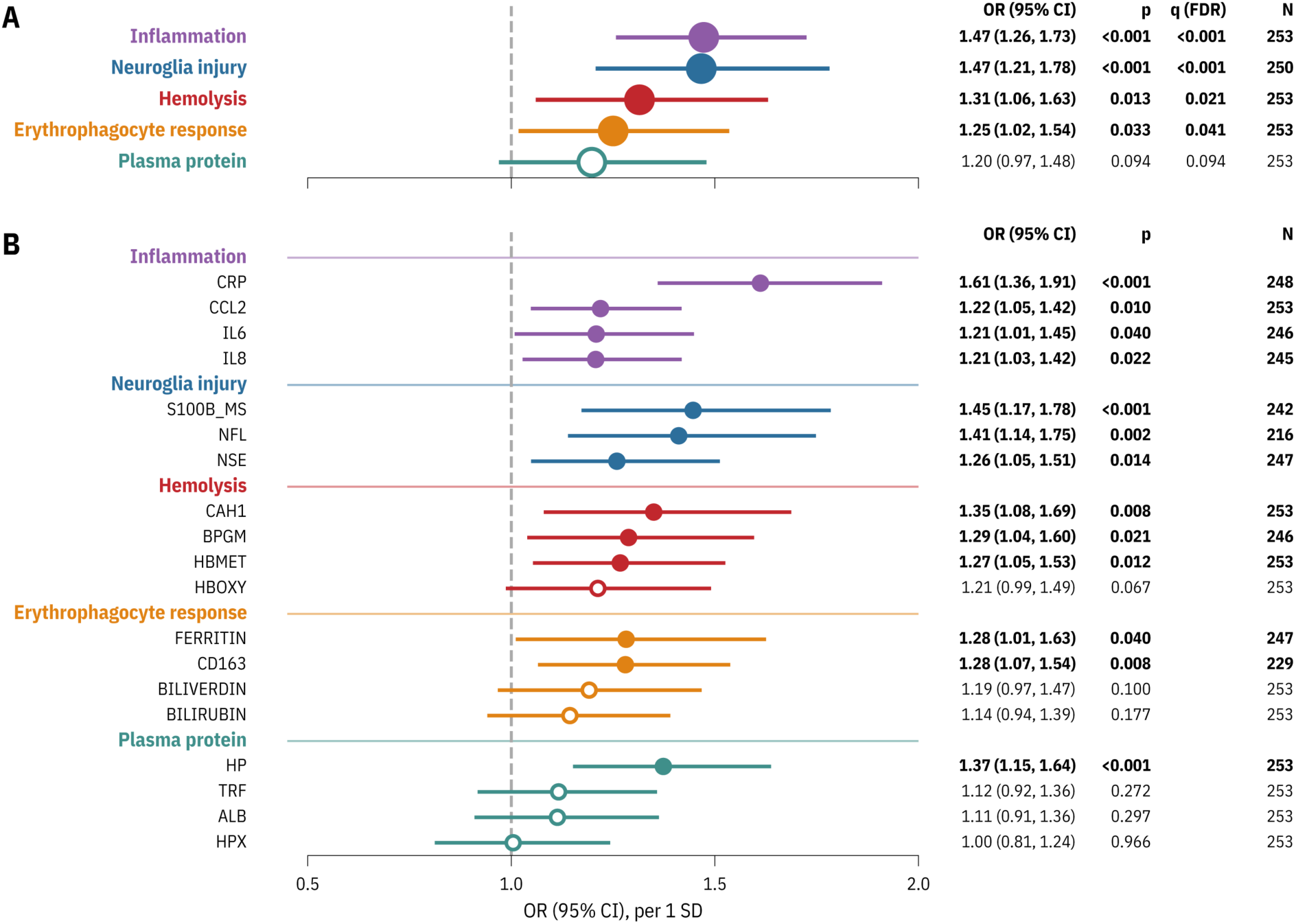
Concurrent same-day association of CSF biomarker pathways and individual biomarkers with secondary brain injury. **(A)** Pathway-level odds ratios for the five prespecified composites from generalized estimating equations (GEE; exchangeable working correlation, categorical day and site fixed effects, robust sandwich standard errors), expressed per 1-SD increase in the pathway score with 95% CIs. The outcome was assessable SBI status on the same day the CSF sample was drawn. Significance was assessed by Benjamini–Hochberg FDR across the five pathways: filled symbols denote q < 0.05. **(B)** Single-biomarker same-day associations for all 19 pathway analytes, grouped by pathway. Estimates are OR per 1-SD increase with 95% CI. Panel B is a descriptive within-pathway visualization; primary inference is at the pathway level in panel A. Filled symbols and the annotated p values are raw, uncorrected p values. Benjamini–Hochberg q values across the 19 analytes were computed and are reported in the deposited result tables but are not displayed here; 11 of the 19 reached q < 0.05, the exceptions among the nominally significant markers being ferritin and IL-6 (both q = 0.059). Working-correlation sensitivity analyses under AR (1) were concordant with the primary pattern (Supplemental Figure 2). Annotation columns give the OR (95% CI), the raw p value, and the number of contributing patients.

Leave-one-marker-out re-estimation preserved the direction of every pathway association, and hemolysis was the most stable composite (all ΔOR within ±3.23%; **Supplemental Table 1, Supplemental Figure 3**). Individual biomarkers moved with their parent pathway in four of the five composites (**Figure 2B**), which supports the composite mean as the primary level of inference. The magnitude of associations showed some variation within pathways, with CRP the dominant contributor to the inflammation composite. Within the plasma-protein composite, which was not associated with same-day SBI, haptoglobin was the only constituent with an individual association (OR 1.37, 95% CI 1.15–1.64), whereas the references albumin and transferrin and the scavenger hemopexin were close to the null. Within the hemolysis pathway, the strongest individual signals were observed for the MRM-quantified erythrocyte proteins CAH1 and BPGM, while the spectrophotometrically measured hemoglobin species showed directionally concordant, but somewhat weaker, associations.

Same-day SBI was therefore associated with higher scores in four pathways: inflammation, neuroglial injury, hemolysis, and the erythrophagocyte response.

### Prospective first-event associations narrow to erythrocyte-derived pathways

To distinguish temporally antecedent pathway activity from concurrent SBI biology, we extended the GEE framework to a first-event horizon (H) analysis. In day-indexed formulas, H denotes days (0, 1, 2, or 3). The H = 0 model tested the association with first SBI. In contrast to the same-day analysis (**Figure 2**), the H = 0 first-event model excluded recurrent-SBI-event days by restricting the analyzed data set to the time period up to and including the day of first SBI. All H > 0 models evaluated the outcome of first SBI on days d+1 through d+H, excluding the sampling day. Benjamini–Hochberg correction across the five composites was applied within every horizon, but only H = 0 and +72 hours were prespecified as inferential; the +24 h and +48 h horizons are shown for continuity and are read descriptively (**Figure 3A**).

**Figure 3.**
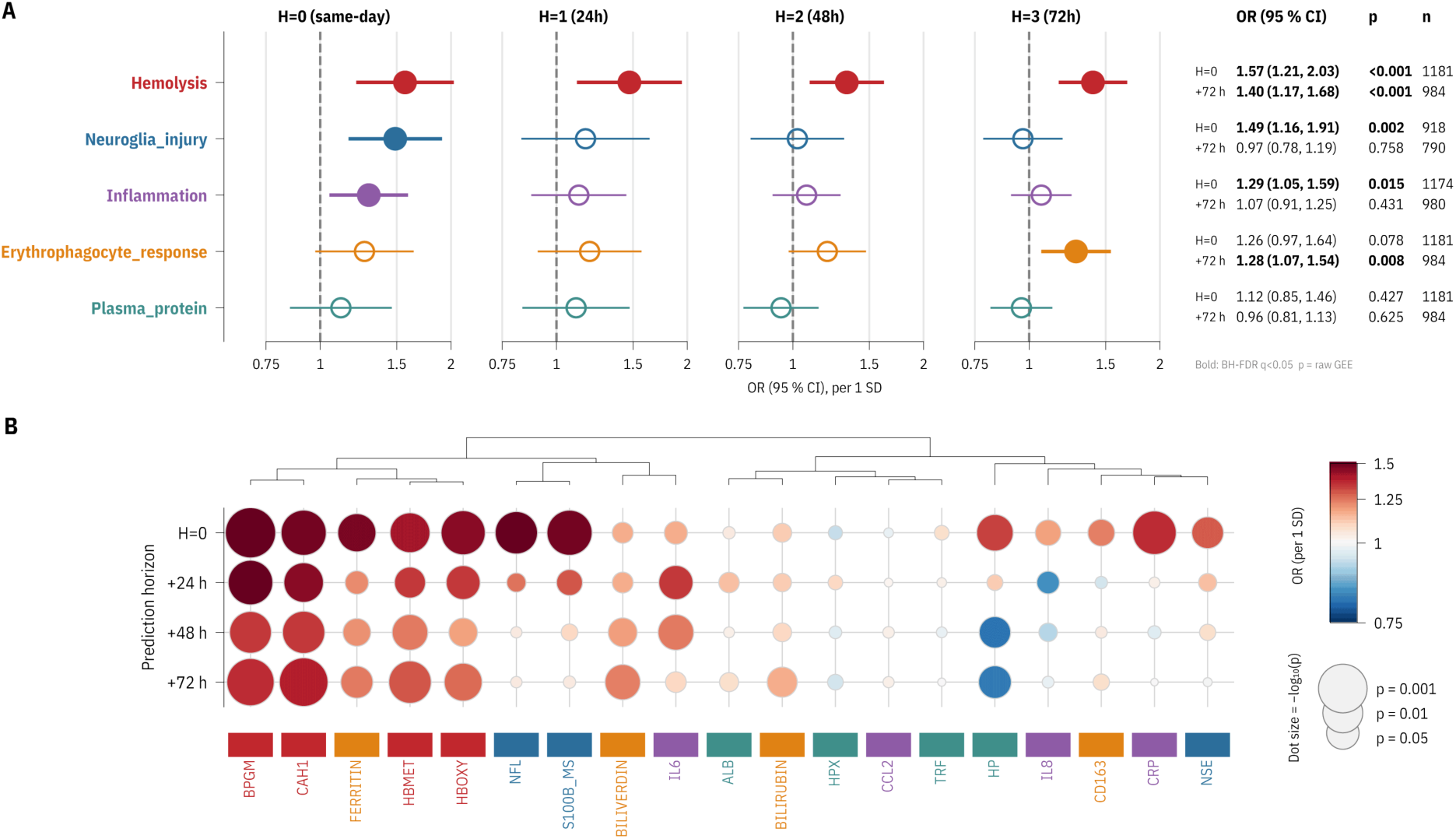
Among patients still event-free, hemolysis and the erythrophagocyte response are the only CSF pathways associated with first secondary brain injury over the next 72 hours. **(A)** Pathway-level odds ratios from horizon GEE models with a cumulative first-event outcome. At H = 0, the outcome was first SBI on the sampling day. At H > 0 (+24, +48, +72-hour), the risk set was event-free through the sampling day and the outcome was first SBI strictly after sampling (see Methods). Models used exchangeable working correlation, categorical day and site fixed effects, and robust sandwich standard errors; estimates are OR per 1-SD increase in the pathway score with 95% CI. Annotation columns give the OR (95% CI), raw GEE p value, and number of contributing patient-days (n); bold OR entries denote Benjamini–Hochberg FDR significance (q < 0.05 across the five pathways), tested at the two prespecified primary horizons (H = 0 and +72-hour); intermediate horizons are shown for continuity. The H = 0 model excludes recurrent-event days and therefore differs slightly from Figure 2A. Hemolysis and the erythrophagocyte response were the only pathways with an FDR-significant prospective +72-hour association; per-pathway estimates and FDR significance are annotated in the panel. **(B)** Secondary biomarker-level characterization of horizon estimates for all 19 pathway analytes, ordered by Ward-linkage hierarchical clustering of the four-horizon log-OR vectors. Dot color encodes OR per 1-SD increase and dot size encodes −log10 (p). Panel B is descriptive; primary inference is at the pathway level in panel A. The clustered biomarker pattern placed all four hemolysis constituents in the high-OR block at the left of the ranking (positions 1, 2, 4 and 5 of 19, interleaved only by ferritin), with the strongest effects for the erythrocyte enzymes CAH1 and BPGM and directionally concordant hemoglobin species.

At the concurrent H = 0 horizon, the first-event analysis was broadly consistent with the same-day model (**Figure 2**). Neuroglial injury, hemolysis, and inflammation remained FDR-significant, whereas the erythrophagocyte response and plasma proteins did not (**Figure 3A**).

As the horizon extended, the pattern narrowed to the erythrocyte-derived pathways. At the prespecified +72-hour horizon, hemolysis remained associated with future first SBI (OR 1.40 per 1-SD, 95% CI 1.17–1.68, q = 0.001), and the erythrophagocyte response was also FDR-significant (OR 1.28 per 1-SD, 95% CI 1.07–1.54, q = 0.021). Inflammation (OR 1.07 per 1-SD, 95% CI 0.91–1.25, q = 0.72), neuroglial injury (OR 0.97, 95% CI 0.78–1.19, q = 0.76) and plasma proteins (OR 0.96, 95% CI 0.81–1.13, q = 0.76) attenuated toward the null and did not reach FDR significance (**Figure 3A**). The neuroglial model included fewer patient-days because of lower constituent-analyte completeness (790 versus 984 for hemolysis). At the intermediate +24 h and +48 h horizons, which were not prespecified as inferential, hemolysis was the only pathway with q < 0.05, and the erythrophagocyte response remained directionally positive throughout.

A secondary biomarker-level analysis (Ward-linkage clustering of the four-horizon log-OR profiles) recapitulated the pathway result: all four hemolysis constituents were directionally concordant and three reached FDR significance at +72 h, strongest for the erythrocyte enzymes BPGM and CAH1 (**Figure 3B**). Within the erythrophagocyte composite no constituent was FDR-significant. Biliverdin was nominally significant at +72 h (raw p = 0.029, q = 0.11) and positive point estimates were observed for biliverdin, bilirubin, and the iron-storage protein ferritin, whereas soluble CD163 was close to the null. Haptoglobin was the only analyte with a nominally significant inverse forward association, higher CSF concentrations tracking with lower odds of subsequent first SBI (+72 h OR 0.82 per 1-SD, 95% CI 0.67–1.00; raw p = 0.049). Four further analytes (hemopexin, IL-8, NSE and CRP) had +72-hour point estimates below 1.0, but all lay within 0.95–1.00 of the null and none approached significance. The haptoglobin estimate did not survive FDR correction (q = 0.15) and is reported descriptively.

Thus, the prospective first-event analysis narrowed the four same-day pathway associations to the two erythrocyte-derived composites, with hemolysis the only pathway reaching q < 0.05 at every horizon examined. At +72 hours, both erythrocyte-derived composites were FDR-significant despite having no analytes in common, and their constituents spanned three analytical platforms. This pattern is consistent with linked stages of erythrocyte lysis and heme processing rather than an association confined to one analyte set or assay, but it does not constitute independent replication.

### Hemolysis precedes SBI in a day-wise landmark analysis

Because both hemolysis and SBI risk rise over the days after hemorrhage, any monotonically increasing analyte can appear to anticipate a later event; the horizon GEE addresses this through categorical day fixed effects. As a complementary, methodologically distinct analysis, we constructed a day-wise landmark GLM analysis in which each comparison is made within a single post-hemorrhage day. The risk set at each landmark day was homogeneous for time since hemorrhage, removing the population trajectory by design rather than by adjustment.

At each landmark day t ∈ {1, …, 11}, the risk set consisted of patients still event-free at day t, and the outcome was first SBI on days t+1 to t+3 (within the +72-hour window). The sampling day t itself was excluded from the outcome. Within each landmark day, sites contributing only events or only non-events were dropped before fitting, since a site without outcome contrast on that day is absorbed by its own fixed effect and cannot inform the estimate. A separate logistic GLM (ridge-penalized for sparse days) was fitted at each landmark day; per-day hemolysis estimates across the evaluated landmark window (days 1–11) are shown in **Figure 4A**.

**Figure 4.**
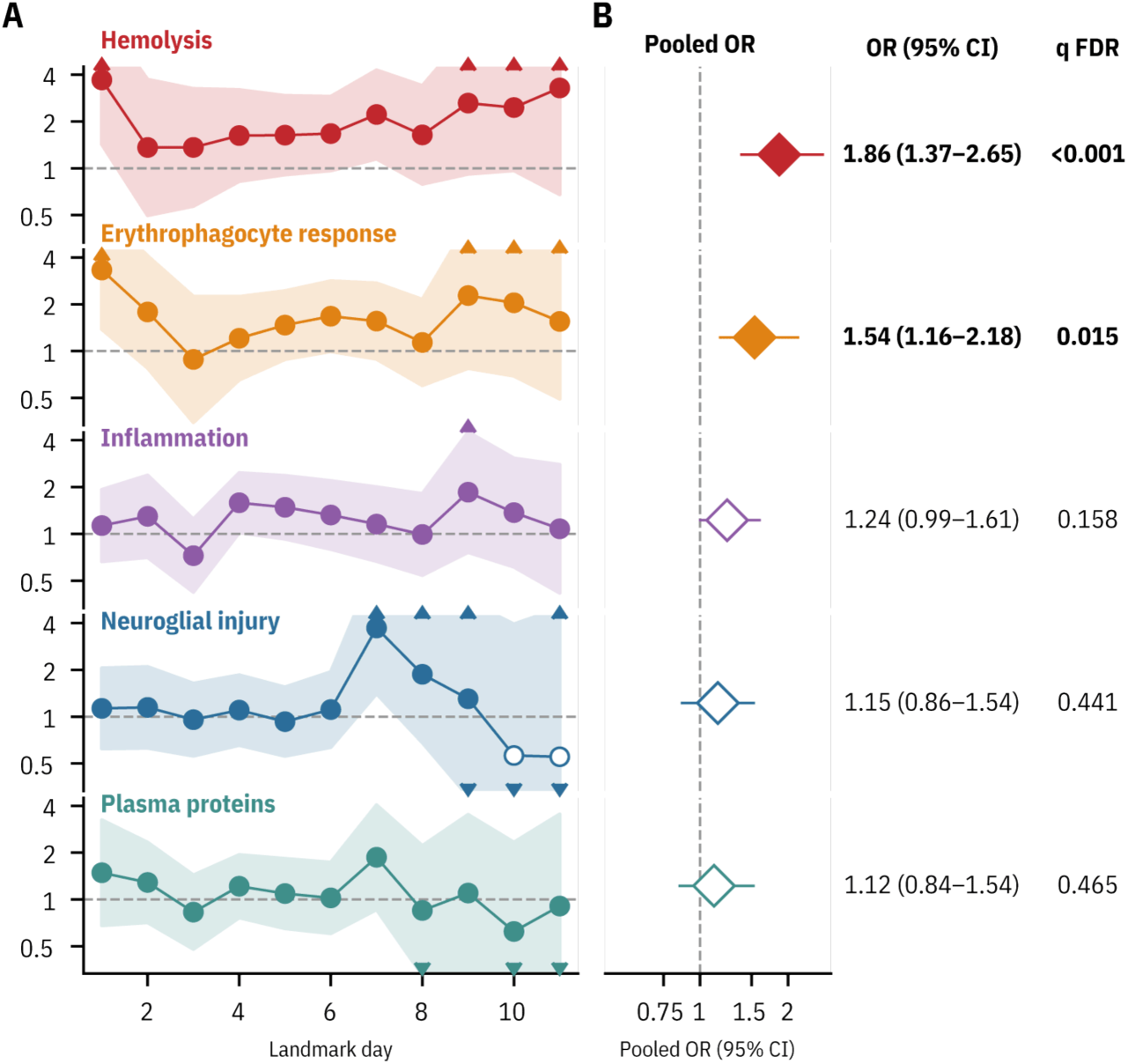
Day-wise landmark GLM analysis of the +72-hour prospective hemolysis signal. **(A)** Per-day landmark odds ratios for SBI within the +72-hour horizon across landmark days 1– 11. Each landmark day used the risk set of patients still event-free at that day and predicted first SBI within the next 72 h (excluding the landmark day itself) from that day’s pathway composite score. Separate logistic generalized linear models (GLMs) were fitted within each landmark day; ridge penalization was used for sparse day-level fits. Empty symbols mark ridge fallback; shaded bands show per-day 95% CIs. **(B)** Per-day log-ORs were pooled by inverse-variance weighting across the unpenalized landmark days carrying a finite Wald-type standard error; ridge-penalized days appear in panel A but did not contribute. All eleven days contributed for every pathway except neuroglial injury, for which days 10 and 11 were ridge-penalized and nine days were pooled. Uncertainty was estimated by patient-level cluster bootstrap (B = 2,000), resampling whole patients to account for cross-day patient overlap. Intervals are bootstrap percentile intervals; the q value is the Benjamini–Hochberg adjustment of the Gaussian bootstrap p value across the five pathways (see Methods). Filled diamonds and bold q values mark q < 0.05; open diamonds are non-significant. Hemolysis (q < 0.001) and the erythrophagocyte response (q = 0.015) were FDR-significant.

Per-day log-ORs were pooled by inverse-variance weighting, with inference from a patient-level cluster bootstrap that resampled patients and rebuilt the full landmark cascade. Hemolysis showed the strongest FDR-significant +72-hour landmark association under this patient-level bootstrap inference (pooled OR 1.86, 95% bootstrap CI 1.37–2.65, q < 0.001; **Figure 4B**), with the erythrophagocyte response also FDR-significant (pooled OR 1.54, 95% bootstrap CI 1.16– 2.18, q = 0.015). All eleven hemolysis landmark days were fitted by standard GLM and contributed to the pooled estimate; ridge penalization was required only for neuroglial injury on days 10 and 11, which are displayed but excluded from pooling. The per-day hemolysis estimates were positive on all eleven landmark days, and the pooled estimate was insensitive to leave-one-day-out removal (OR range 1.78–1.91). Inflammation, neuroglial injury and plasma proteins did not reach FDR significance. Inflammation remained positive but imprecisely estimated; its interval included both the null and moderate positive associations.

Both analyses identified hemolysis and the erythrophagocyte response at +72 hours, with a larger point estimate for hemolysis in each. Because they used the same parent risk set, exposure, and outcome, their concordance supports robustness to model specification rather than independent replication. The horizon GEE estimated a joint day-adjusted repeated-measures association, whereas the landmark analysis estimated and pooled within-day associations using patient-level cluster-bootstrap inference.

### Event-centered trajectories show pre-SBI rises in erythrocyte-derived pathway scores

We next asked whether the prospective hemolysis pattern was visible descriptively in patients who experienced SBI, re-centering pathway scores on each patient’s first-SBI day (**Figure 5**). Non-assessable days were retained as biomarker observations that contributed to patient trajectory but not to event timing.

**Figure 5.**
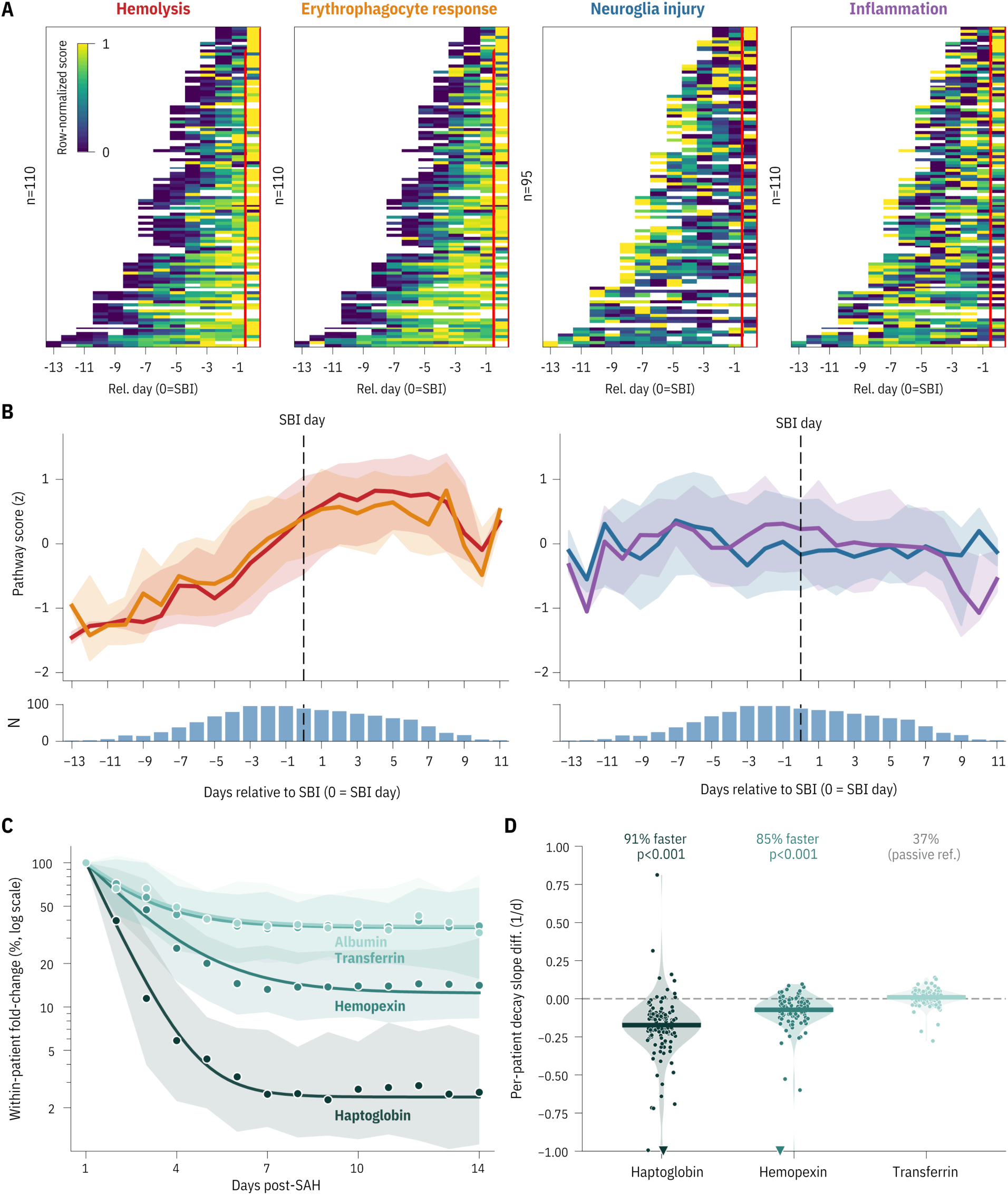
Event-centered CSF pathway scores and selective scavenger-protein depletion after aSAH. **(A)** Per-patient pathway scores were aligned to each patient’s clinically documented first SBI day, defined as day 0; this is the event day recorded in the clinical database, not the first sampling day on which SBI was captured. Non-assessable days were retained for descriptive biomarker trajectories. Columns show days relative to first SBI, up to and including day 0, and rows are sorted by first-SBI day. Values are within-patient min–max normalized to a 0–1 scale across the displayed window. Hemolysis and the erythrophagocyte response show visible pre-event build-up, whereas neuroglial injury and inflammation show less consistent pre-event rise. The plasma-protein composite is not shown here; its constituents are characterized in C–D. Of the 154 patients with a documented first SBI day, 144 contributed at least one sampled day on or before it; a patient enters a given panel only if at least three of those days carry a non-missing score for that pathway, which yields n = 110 (hemolysis), 110 (erythrophagocyte response), 95 (neuroglial injury) and 110 (inflammation). The neuroglial panel is smaller because its composite draws on only three constituents, the least completely measured of the five. **(B)** Median pathway score with interquartile range over days relative to first SBI, plotted in the original z units of the composite and not min–max normalized. Bar plots show the number of contributing patients per relative day. The dashed vertical line marks the first SBI. Panels A–B are descriptive event-centered displays and did not contribute to formal inference; the scavenger analysis in C–D is independent of SBI status. **(C)** Descriptive scavenger-protein depletion analysis in 131 patients with an available day-1 EVD-CSF sample. Within-patient fold-change trajectories are shown relative to each patient’s own day-1 concentration for haptoglobin, hemopexin, albumin, and transferrin. Points show daily medians, shaded bands indicate interquartile ranges, and smooth curves summarize descriptive monoexponential decay-to-plateau fits. Haptoglobin and hemopexin were depleted to lower residual plateaus than albumin and transferrin. **(D)** Per-patient log-decay-slope differences relative to albumin, restricted to patients with an estimable slope for both proteins in the pair (at least three positive fold-change measurements on at least two distinct days): n = 122 for haptoglobin and transferrin, 121 for hemopexin, of the 131 patients in C. Values below zero indicate faster decline than albumin. Haptoglobin and hemopexin declined faster than albumin in 91% and 85% of these patients, respectively, whereas transferrin showed no scavenger-like shift toward faster decline. Groups were compared with the Wilcoxon signed-rank test.

Both per-patient intensity heatmaps (**Figure 5A**) and median trajectories (**Figure 5B**) showed a progressive pre-event rise in hemolysis and the erythrophagocyte response, whereas inflammation and neuroglial injury stayed near mean levels until at or after the event. This descriptive visualization was concordant with the prospective models and did not contribute to formal inference.

### Haptoglobin and hemopexin are selectively depleted from CSF

To compare endogenous scavenger trajectories with those of reference plasma proteins, we descriptively examined constituent trajectories in 131 patients with an EVD-CSF sample available on day 1 (**Figure 5C**). After within-patient normalization to each patient’s own day-1 level, albumin and transferrin showed similar declining profiles and residual plateaus, whereas haptoglobin and hemopexin were disproportionately depleted. Over days 1–14, CSF haptoglobin declined to a residual plateau of 2.4% of each patient’s day-1 concentration (bootstrap 95% CI, 1.6–3.4) and hemopexin to 12.5% (10.3–15.9), while the passive plasma-protein references albumin and transferrin plateaued at 35.5% (27.3–41.8) and 36.1% (30.2– 46.0). As a model-free check, the raw day-7 haptoglobin value (∼2.5% of day-1) closely matched the fitted plateau and was near or below the lower limit of quantification, compared with approximately 35% for albumin.

Paired within-patient slope analyses confirmed that haptoglobin and hemopexin declined faster than albumin in most patients (111 of 122 slope-eligible patients, 91%, for haptoglobin and 103 of 121, 85%, for hemopexin; Wilcoxon signed-rank p < 10⁻¹⁵ for both, **Figure 5D**). Transferrin, a second passive plasma-protein reference, was centered near albumin and did not show a scavenger-like shift toward faster decline. Despite late haptoglobin censoring below the assay lower limit of quantification, the concordant hemopexin pattern and passive-reference comparison support selective scavenger consumption or clearance rather than passive plasma-protein washout alone.

## DISCUSSION

In this prospective multicenter analysis of serial CSF after aSAH, inflammation and neuroglial injury showed the largest same-day associations with SBI, whereas only hemolysis and the linked erythrophagocyte response remained FDR-significant at the prespecified +72-hour first-SBI-event horizon. Hemolysis was the strongest prospective signal across analyses, was carried most strongly by redox-state-independent erythrocyte-lysis markers rather than by spectrophotometric hemoglobin species, and occurred in a compartment where haptoglobin and hemopexin were selectively depleted. Parallel assessment of five pathways within the same cohort compares biological signals before first SBI with those accompanying its clinical recognition.

Our findings both confirm and reframe prior CSF biomarker work in aSAH. Earlier work, particularly studies focused on inflammatory mediators, has consistently linked cytokines and related markers to delayed injury and poor outcome(31–35). Neuroglial injury markers have also been found to be associated with early and secondary brain injury after aSAH(36, 37). HeMoVal is consistent with these observations in a large prospective multicenter CSF cohort. The distinction our design adds is timing. A same-day association links biomarker levels to SBI documented on the sampling day. The forward models use samples collected while patients remained event-free. In the +72-hour GEE analysis, inflammation and neuroglial injury did not retain FDR-significant associations with first SBI within the next 3 days. By contrast, hemolysis remained associated with later SBI across analytical approaches. This pattern supports blood-product toxicity as a candidate upstream process(26). The day-wise landmark analysis, which compares patients within the same post-hemorrhage day, further separates this pattern from a shared global time-trend.

The biomarker-level results provide biological context for the negative primary HeMoVal monitoring result. A single labile analyte can fail as a bedside monitor while the broader biological pathway it indexes remains informative. The prospective hemolysis association was carried most strongly by the MRM-quantified erythrocyte proteins BPGM and CAH1, with directionally concordant hemoglobin species. This is biologically coherent. The spectrophotometric hemoglobin species are redox-state-dependent, HbOxy oxidizes to HbMet, and oxidized hemoglobin degrades with heme release(38, 39), whereas BPGM and CAH1 are redox-state-independent markers of erythrocyte lysis. Consistent with this, the companion monitoring analysis found that among spectrophotometric species it was the oxidized metabolite methemoglobin that remained associated with same-day SBI, in a post-hoc exploratory analysis(25). The present data place that same-day spectrophotometric signal within a broader erythrocyte-lysis signature associated with subsequent SBI. The study-specific mass-spectrometry panel was therefore an important enabler of the pathway-level analysis, allowing robust quantification of erythrocyte, scavenger, neuroglial, and reference proteins in the same CSF samples.

The temporal pattern observed in HeMoVal aligns with prior experimental and human evidence (**Table 1**). Experimental work shows that extracellular hemoglobin disrupts nitric oxide-sensitive vascular signaling(7, 40) and that haptoglobin can attenuate hemoglobin-induced vasospasm and the associated oxidative, glial, and neuronal injury(16–19, 41, 42). Human imaging, prior CSF cohorts, and a Mendelian randomization study also implicate hemolysis and scavenger protein biology as potential determinants of aSAH outcome(6, 20–22, 43, 44). Earlier CSF studies linked hemoglobin to subsequent neurofilament light, and untargeted proteomics has related day-7 profiles to six-month functional outcome after aSAH (21, 45). HeMoVal extends this evidence through dense multicenter sampling and comparisons of hemolysis-related, inflammatory, and neuroglial pathways prior to the first documented SBI.

**Table 1.** Triangulation of evidence. Distinct evidence streams support hemoglobin toxicity and scavenger protection through different designs and biases.

| Evidence stream (design) | Main contribution | Limitation addressed by HeMoVal |
| --- | --- | --- |
| Experimental Hb/heme toxicity in organotypic brain slices (*in vitro*), rodents (*in vivo*), and isolated cerebral arteries (*ex vivo*) | Cell-free Hb depletes nitric oxide, impairing NO-sensitive vascular signaling; Hb and heme drive oxidative stress, iron accumulation, and glial and neuronal injury; haptoglobin and hemopexin attenuate toxicity in these controlled systems. | Controlled exposure models cannot establish complex endogenous pathway timing before and during SBI |
| Large-animal translational exposure and rescue studies in sheep | Acute CSF-Hb injection causes cerebral vasoconstriction. Repeated CSF-Hb injection causes neurological deterioration over 3 days; haptoglobin attenuates these phenotypes. | Experimental Hb administration does not reproduce the endogenous human disease course. |
| Prior human CSF studies: small single-center serial-CSF cohorts and ex vivo assays with patient CSF | CSF-Hb tracks SBI and predicts subsequent CSF neurofilament light neuronal-injury marker; Hb-rich patient CSF causes ex vivo cerebral artery vasospasm and lipid peroxidation, attenuated by haptoglobin and hemopexin. | Limited sample sizes and differing endpoints; no multicenter comparison of CSF pathways before first documented clinical SBI. |
| Mendelian randomization (genetic instrument for CSF haptoglobin) | Higher genetically predicted CSF haptoglobin associates with lower catastrophic aSAH outcome, but not lower aSAH occurrence. | Genetic instruments do not resolve when CSF pathways are active relative to SBI. |
| **HeMoVal** (present study)<br>Prospective multicenter serial-CSF cohort with comparative disease pathway analysis | Hemolysis and its downstream erythrophagocyte response are the only prespecified CSF pathways with an FDR-significant +72-hour first-event association, while the concurrently dominant inflammation and neuroglial injury did not retain FDR-significant associations at +72 hours. | Adds a multicenter, temporally resolved human comparison in which hemolysis and the erythrophagocyte-response composite retained FDR-significant associations with first SBI within the next 72 hours. |

Albumin and transferrin concentrations declined similarly, whereas haptoglobin and hemopexin declined more rapidly. The greater hemopexin decline despite molecular size comparable to albumin and transferrin, which declined similarly, is not readily explained by size-dependent clearance alone. This pattern is consistent with hemoglobin and heme exposure occurring in a CSF compartment with progressively diminishing endogenous scavenger capacity(46, 47).

Haptoglobin was positively associated with same-day SBI but showed a nominal, non–FDR-significant inverse forward association. This sign difference is compatible with, but does not establish, remaining buffering capacity. If hemolysis contributes upstream to SBI, the pre-event hemolysis association and progressive scavenger decline support testing intrathecal haptoglobin supplementation to sustain CSF hemoglobin scavenging across the risk period(26). Similarly, strategies that accelerate clearance of blood products from the CSF, such as early or augmented CSF drainage(48), may act on the same upstream hemolysis axis.

Several features strengthen the interpretation of these findings, including prospective multicenter sampling, standardized prospective SBI assessment, locked clinical databases before biomarker analysis, dense EVD-CSF collection, and a prospectively assembled multi-platform biomarker panel. Initial hemorrhage burden or clinical severity could partly explain the hemolysis association. Hemorrhage burden is also a source of the measured hemolysis exposure and was associated with CSF cell-free hemoglobin in the primary report(25). The attenuation of inflammation and neuroglial injury at +72 hours, together with persistence of both erythrocyte-derived composites, argues against a wholly nonspecific severity signal but does not exclude severity-related confounding.

Important limitations remain. First, this observational study establishes antecedence but not causation, and residual confounding, including severity-related structure, cannot be fully excluded. Second, no formal contrasts between pathway coefficients or across horizons were performed. Differences in point estimates or FDR status therefore describe the pattern of separately estimated associations and should not be interpreted as tested differences between effects. Third, because the cohort was restricted to patients requiring EVD, generalizability to milder aSAH is limited. Fourth, the registered SBI composite aggregates related but biologically non-identical states (angiographic vasospasm, delayed cerebral ischemia, and delayed ischemic neurological deficit), which may blur pathway-specific timing and dilute associations. Fifth, ventricular EVD-CSF is an imperfect proxy for the subarachnoid space in which much of SBI evolves. Biomarker concentrations in EVD-CSF are affected by dilution from continuous CSF production and drainage, variable blood contamination, and CSF-diversion dynamics. Many of these sources of measurement noise would be expected to attenuate associations.

Sixth, biomarker missingness was largely assay-dependent and was mitigated by the use of pathway composites, which require only two of the constituent analytes, but complete-case modeling remains a limitation and the missingness mechanism was not formally tested. Seventh, the cumulative horizon design trades sample for reach: as the horizon lengthens, the eligible risk set contracts (1,318, 1,162, 1,119 and 1,068 patient-days at H = 0, +24, +48 and +72 hours, before predictor-specific missingness and stratum pruning) and is truncated to progressively earlier post-hemorrhage days, so the more distal estimates are drawn from a narrower and earlier slice of the sampling window. The count of positive outcome windows, by contrast, rises across the forward horizons (77, 155 and 230 at +24, +48 and +72 hours). These are not independent events: a wider window captures the same first SBI from more preceding sampling days, so the increase reflects longer look-forward rather than additional patients experiencing SBI. Eighth, patients with no SBI-positive EVD-CSF sampling day contributed fewer sampling days, so the at-risk sampling is subject to potential informative censoring. The first-event and day-wise landmark designs do not eliminate potential bias from informative sampling, censoring, or death before SBI ascertainment. Ninth, although the biomarker panel, the five-pathway architecture, and our research program’s focus on hemolysis biology were prespecified, the specific finding that erythrocyte-derived pathways would be the *only* ones with a forward-significant association, dissociating from the concurrently dominant inflammatory and neuroglial pathways, was not stated as an a priori hypothesis.

In patients with aSAH, hemolysis within the CSF compartment precedes secondary brain injury. The hemolysis signal is carried most strongly by redox-stable erythrocyte-lysis markers rather than a single hemoglobin species measured by spectrophotometry, and unfolds in a compartment where endogenous scavenger proteins are selectively depleted. Together with prior experimental and human evidence, these findings support hemolysis as a candidate upstream pathobiological process in aSAH, and provide a rationale for testing sustained hemoglobin-scavenging strategies within the CSF compartment.

## METHODS

### Sex as a biological variable

Both sexes were included. Consistent with aSAH epidemiology(49), the EVD cohort was predominantly female (68.3%; n = 177/259). The underlying primary study was not designed to estimate sex-specific effects, and sex was not included as a covariate in the association models.

### Study design and patient population

HeMoVal (Hemoglobin Monitoring after Subarachnoid Hemorrhage Validation) was a preregistered, prospective, international multicenter observational cohort study (ClinicalTrials.gov NCT04998370) conducted at eight academic neurosurgical centers in Switzerland, Germany, and Austria. The study was registered before recruitment, the protocol was subsequently published open access, and the primary monitoring analyses followed a prespecified statistical analysis plan(24). Adults (≥18 years) with radiologically confirmed aneurysmal SAH were screened within 24 h of admission; non-aneurysmal SAH, concurrent enrollment in another interventional or CSF-sampling study, and prior HeMoVal enrollment were exclusion criteria. The registered primary monitoring objective and the full enrolled cohort have been recently reported(25). The biomarker pathway analyses reported here were mandated by the same published protocol, which prospectively specified daily CSF biobanking and biomarker-based pathway analyses; the present work is therefore a distinct, prespecified secondary analysis of the HeMoVal CSF biobank rather than an overlapping report of the registered primary monitoring endpoint.

The present analysis used the EVD-CSF cohort. CSF (2 mL) was sampled once daily from day 1 to day 14 after hemorrhage whenever an external ventricular drain was in place and sampling was not contraindicated, centrifuged immediately (1,500 ×*g*, 15 min), and the supernatant stored at −80 °C as part of the prospectively mandated HeMoVal biobank. SBI was assessed daily over the 14-day window as a composite of angiographic vasospasm, delayed cerebral ischemia, and delayed ischemic neurological deficit, according to the standardized, published protocol(24). Clinical investigators were blinded to biomarker results, and the clinical databases were locked before biomarker measurements were performed and before the clinical and biomarker databases were merged for inferential analysis. The biomarker analysis comprised 259 patients with EVD-sampled CSF contributing 2,411 valid-day; 1,925 patient-days from 253 patients with paired biomarker measurements and assessable SBI status entered inferential models, whereas 486 non-assessable patient-days were retained for descriptive trajectory displays only.

### CSF biomarker quantification

CSF biomarkers were quantified across three complementary analytical platforms. Hemoglobin species (oxyhemoglobin, methemoglobin) and the heme catabolites bilirubin and biliverdin were quantified using UV/Vis spectrophotometry (Lunatic, Unchained Labs; 350–750 nm) with spectral deconvolution as previously described (25). Cytokines and cellular markers (CRP, IL-6, IL-8, CCL2, C5a, syndecan-1, CD163, ferritin, NSE, D-dimer) were measured by immunoassay using multiplex panels from R&D Systems. All assays were conducted in accordance with the manufacturers’ protocols. Eight plasma, erythrocyte, scavenger, and neuroglial proteins [albumin, transferrin, haptoglobin, hemopexin, carbonic anhydrase 1 (gene *CA1*; UniProt P00915, abbreviated CAH1 here), bisphosphoglycerate mutase (BPGM), neurofilament light chain (NfL), and S100B] were quantified by an 8-plex multiple-reaction-monitoring (MRM) liquid chromatography-mass spectrometry assay developed specifically for HeMoVal.

The MRM assay provides absolute quantification of each protein from a 10-µL CSF aliquot via one or two proteotypic surrogate peptides released by tryptic digestion, with stable-isotope-labeled forms of every surrogate peptide added before digestion as internal standards.

Quantification used the peak-area ratio of the endogenous (light) surrogate peptide to its heavy internal standard, interpolated on matrix-matched calibration curves. Artificial CSF was used for spiking of calibration standards, and quality control samples at different spike levels were used to validate the adequacy of the surrogate matrix in quantifying endogenous human proteins in the clinical samples. The assay was qualified in accordance with ICH Q2 (R1)(50) and the FDA Bioanalytical Method Validation guidance and met all predefined acceptance criteria for accuracy, intra-and inter-assay precision, linearity (r² ≥ 0.985 across each analyte’s validated range), lower and upper limits of quantification, specificity, dilution integrity, on-instrument stability, and carryover (Supplemental Methods; Supplemental Tables 2 and 3). Because post-hemorrhagic CSF is itself a hemolytic matrix, we spiked pooled human CSF with 0.5%, 1%, and 2% hemolyzed whole blood; recovery of all eight quantifier peptides stayed within ±15% of nominal. Of the MRM analytes, CAH1 and BPGM are erythrocyte-enriched enzymes released during red-cell lysis and entered the hemolysis composite; NfL and S100B entered neuroglial injury; and albumin, transferrin, haptoglobin, and hemopexin constituted the plasma-protein composite. Full reagent sources, peptide and transition lists, sample-preparation and chromatographic conditions, and complete validation results are provided in the Supplemental Methods and Supplemental Tables 2 and 3.

### Statistical analysis

#### Biomarker processing and pathway composites

Each measured CSF analyte was standardized across all EVD CSF samples by a skew-aware z-transform: a Yeo–Johnson power transform(51) was applied when the analyte was substantially skewed, followed by centering and scaling, with standardized values clipped at ±8 to limit leverage. Missing values were left missing and not imputed, as pathway composites could be calculated from available constituent analytes and imputation would require additional assumptions about the underlying biomarker distributions and missingness mechanism. Two analytes fell below the prespecified 50% valid-measurement threshold (C5a, syndecan-1) and D-dimer had no pathway assignment; all three were dropped from the composites and from the inferential panels, but remain in the deposited dataset. This left 19 analytes in five composites: hemolysis (4), erythrophagocyte response (4), inflammation (4), neuroglial injury (3), and plasma proteins (4). Each composite score was the row-wise mean of its available constituent z-scores (requiring ≥2 non-missing constituents) and was re-standardized to unit SD, so that odds ratios are expressed per 1-SD increment in the pathway score. Standardization was performed once per analysis frame and not repeated within individual models: once within the 1,925-row inferential parent set, which supplies the predictor for the same-day, first-event and landmark analyses, and separately within the 2,411-row descriptive set used for the temporal and event-centered displays. A 1-SD increment therefore has the same meaning throughout **Figures 2–4**. The event-centered panels both start from the 2,411-row composite but display it differently: **Figure 5B** retains the composite’s z units as standardized in that descriptive frame, whereas **Figure 5A** rescales each patient’s own trajectory to a 0–1 range and so displays no z scale at all. Neither is numerically interchangeable with the model estimates. The pathway trajectories in **Figure 1F** are constructed differently again: they show the daily median of each constituent analyte’s z-score, with the bold line the unweighted mean of those per-analyte medians, and therefore do not correspond to either composite scaling. Biomarker missingness was characterized descriptively as per-analyte availability and the per-row distribution of available constituents.

#### Concurrent (same-day) associations

We estimated the association between a pathway’s same-day CSF level and the odds of concurrent SBI on a given patient-day, holding day and site constant. This primary concurrent analysis related each pathway composite to same-day assessable SBI status using a logistic generalized estimating equations (GEE) model(52) with an exchangeable working correlation by patient, robust sandwich standard errors, and categorical day and site fixed effects. An exchangeable working correlation was selected for the primary analysis because AR (1) models showed more frequent convergence failures, particularly in sparse outcome strata, whereas the exchangeable specification provided more stable estimates across pathways and biomarkers. Day was entered as categorical levels with automatic coarsening on sparse strata to avoid perfect separation, and the finest separable scheme was retained. Robustness to the assumed within-patient dependence was assessed under an AR (1) working correlation. Descriptively, each individual biomarker was analyzed in the same way.

#### Prospective first-event associations

To separate antecedent from concurrent biology, the GEE framework was extended to a first-event risk set (all post-first-event rows excluded) with horizon-specific outcome definitions. At H = 0 the outcome was first SBI on the sampling day. For all H > 0 horizons (+24, +48, +72 hours) the sampling day was excluded from the outcome: a patient-day d contributed only if the patient was event-free through day d, and the outcome was first SBI occurring strictly after sampling, on days d+1 through d+H; thus no same-day first-SBI row contributed to the +24-, +48-, or +72-hour models. Censoring followed the rule *Y = 1 or d + H ≤ last assessed day*: a patient-day on which first SBI occurred within the horizon remained eligible irrespective of later follow-up, whereas a non-event patient-day was eligible only if the patient was assessed through day d+H. Patient-days later than day 14 − H were additionally excluded, because no non-event outcome could be observed within the ascertainment window. Models retained the exchangeable working correlation, categorical day and site fixed effects, and robust standard errors. After predictor-specific missingness exclusions, day and site strata carrying no outcome contrast were dropped before fitting, since a stratum that is entirely event or entirely non-event is absorbed by its own fixed effect and cannot inform the exposure coefficient; site E, which contributed no first SBI within the analyzed windows, was removed at every horizon. H = 0 and +72-hour were the prespecified primary horizons; intermediate horizons were reported descriptively for continuity. As a secondary view, all 19 analytes were ranked by Ward-linkage hierarchical clustering of their four-horizon log-OR vectors; this ordering is descriptive and was not subjected to formal inference.

#### Day-wise landmark analysis

As a complementary analysis, a day-wise landmark analysis(53) fitted a separate logistic model at each landmark day t (risk set = patients still event-free at day t; outcome = first SBI within (t, t + 72 h]), with L2-ridge penalization as a fallback for sparse day-level fits. Per-day log-odds ratios from days with finite Wald-type standard errors were pooled by inverse-variance weighting (ridge-penalized sparse days were displayed but did not contribute to the pooled estimate), and inference was based on a patient-level cluster bootstrap (B = 2,000 resamples; fixed seed) that resampled whole patients, rebuilt the full landmark cascade, and re-pooled across days. Confidence intervals are bootstrap percentile intervals. The p value carried forward to FDR correction was the Gaussian bootstrap p, 2Φ (−|β^|/SE_boot), computed from the bootstrap standard error. The percentile bootstrap p is bounded below by 1/B (0.0005 here), a bound reached by hemolysis, so Benjamini–Hochberg adjustment of the percentile p values assigns hemolysis and the erythrophagocyte response a common q of 0.0025 and cannot separate them; the percentile p is therefore retained as a diagnostic only. The naive independence estimate is reported alongside for comparison.

#### Multiple testing, significance, and descriptive analyses

Pathway-level inference was the primary analysis level. Benjamini–Hochberg false-discovery-rate (FDR)(54) control was applied across the five pathways — within each horizon for the horizon GEE, and across pathways for the same-day and landmark analyses. Only H = 0 and +72 hours were prespecified as inferential; q values at the intermediate horizons were computed on the same basis but are read descriptively. Biomarker-level analyses were secondary: FDR-adjusted q values were computed across the 19 analytes and are reported in the deposited result tables, but interpretation is anchored at the pathway level. A two-sided FDR q < 0.05 was considered significant.

Descriptive temporal structure was summarized with event-centered pathway trajectories re-aligned to each patient’s first-SBI day. Analyses were performed in Python (numpy, pandas, statsmodels, scikit-learn, scipy, matplotlib); stochastic procedures used a single fixed master seed (202501) throughout. Full specifications are provided in the Supplemental Methods.

#### Model diagnostics and quality control

Assay performance was analytically qualified. For the regression models, fit was examined using per-pathway diagnostic grids of Normal Q–Q plots of randomized quantile residuals, calibration curves, and binned residual-structure plots, generated for the concurrent same-day GEE (**Supplemental Figure 1**) and, in the deposited results, for the prospective first-event models at H = 0 and +72 hours. These diagnostics did not indicate misspecification that altered the pathway-level conclusions: calibration and randomized quantile residual plots were consistent with adequate fit in the concurrent and H = 0 models, with a mild shared calibration attenuation at the +72-hour horizon that did not affect the relative pathway ranking. We further assessed robustness with the sensitivity checks the analysis supports: concordance of the concurrent GEE under an AR (1) versus exchangeable working correlation; leave-one-marker-out reconstruction of every pathway composite; stability of the landmark pooled estimate under a patient-level cluster bootstrap and leave-one-day-out recomputation; and automatic day-level coarsening to prevent perfect separation. Estimates were stable across these checks. Because the site-pooling criterion was never met in this cohort (see Supplemental Methods), the primary horizon models coincide with a no-site-pooling specification by construction rather than by separate refitting.

#### Descriptive scavenger-protein depletion analysis

As a supportive descriptive analysis, independent of SBI status and outside the pathway-level FDR inference, we examined EVD-CSF trajectories of the endogenous scavengers haptoglobin and hemopexin together with the passive plasma-protein references albumin and transferrin over days 1–14 after hemorrhage. Analyses were restricted to patients with an available day-1 sample, enabling within-patient normalization of each analyte to that patient’s own day-1 concentration. Daily fold-change trajectories were summarized by median and interquartile range. To summarize trajectory shape, median fold-change curves were fitted with a descriptive monoexponential decay-to-plateau function, FC(t) = P + (1 − P) exp[−k(t − 1)], yielding a residual plateau and apparent half-life; these fits were not interpreted as mechanistic compartmental models. Uncertainty in each residual plateau was quantified by a patient-level cluster bootstrap (B = 2,000 resamples; fixed seed 202501) that resampled whole patients and refitted the decay-to-plateau function to each replicate’s per-day median fold-change, yielding percentile 95% confidence intervals. To compare scavenger depletion with passive plasma-protein behavior, patient-specific log-fold-change slopes were estimated for proteins with at least three positive measurements across at least two days. Slopes for haptoglobin, hemopexin, and transferrin were compared within patients with the albumin slope, and paired differences were tested using Wilcoxon signed-rank tests.

#### Reporting

The study is reported in accordance with the STROBE guidance for observational cohort studies(55); a completed STROBE checklist is provided as a supplementary file. As this is a prespecified secondary pathobiology analysis rather than a diagnostic/prognostic accuracy study, no predictive-performance metric is derived. Full baseline demographic and severity characteristics of the enrolled cohort are reported in the registered primary analysis(25); analysis-set and serial-sampling characteristics specific to this biomarker study are summarized in Supplemental Tables 4 and 5.

#### Study approval

The HeMoVal study was approved by the responsible research ethics committee at each participating center and conducted in accordance with the Declaration of Helsinki and ICH–GCP guidelines; written informed consent was obtained from patients or their legal representatives(25).

#### Data and code availability

De-identified analysis data and the full analysis code are available via Zenodo (dataset: https://doi.org/10.5281/zenodo.20733070; code: https://doi.org/10.5281/zenodo.20733246).

## Supporting information

STROBE checklist

## Data Availability

https://doi.org/10.5281/zenodo.20733246

## ACKNOWLEDGMENTS

We thank the members of the HeMoVal study group and we are grateful to the patients for their participation in and commitment to this study.

## AUTHOR CONTRIBUTIONS

*Conceptualization:* RMB, DJS. *Methodology:* SW, SMP, KA, FV, DC, AS, TG. *Formal analysis:* RMB, DB, UH, DJS. *Investigation:* HeMoVal study group. *Resources:* RMB, KA, MH, DJS. *Data curation:* MS, DC, JF, AW, MI. *Writing – original draft:* RMB, FV, DJS. *Writing – review & editing:* all authors. *Supervision:* UH, TG, DJS. *Project administration:* RMB, KA, MS, TG, DJS.

## AI DISCLOSURE

Generative AI tools (OpenAI GPT family up to GPT-6 Astra and Anthropic Claude family up to Opus-5) were used for language editing, manuscript organization, coding support, and consistency checking against author-verified result files. AI tools did not generate study data or execute the locked statistical analyses. The authors critically reviewed all output, made the final scientific interpretations, and assume full responsibility for the integrity and accuracy of all content.

## Author list order determination description

The order of the co–first authors was determined based on their relative and complementary contributions to the study. RMB led study conceptualization, formal analysis, and preparation of the original manuscript and made leading contributions to the underlying clinical study and cohort framework. KA made leading contributions to the design, establishment, and clinical framework of the underlying cohort and study. SW made leading contributions to the biomarker methodology. SMP developed and analytically validated the study-specific multiplex multiple-reaction-monitoring mass-spectrometry assay for biomarker quantification.

## COLLABORATORS

### HeMoVal study group (as of 31 August 2026)

Amr Abdulazim

Kevin Akeret

Carolin Albrecht

Linda Bättig

Kathrin Susann

Bieri Raphael M. Buzzi

Gwendoline Canzanella-Boillat Elisa Colombo

Daniel Couto

Nima Etminan

Thomas Gentinetta

Matthias Gmeiner

Alexandra Grob

Andreas Gruber

Basil E. Grüter

Joshua Haegler

Ulrike Held

Muriel Helmers

Isabel C. Hostettler

Michael Hugelshofer

Marlies Illi

Yasin Irmak

Vincens Kaelin

Emanuela Keller

David Kronthaler

Sarina Mahler

Bernhard Meyer

Luca Regli

Constantin Roder

Karl Roessler

Moritz Saxenhofer

Dominik J. Schaer

Nina Schwendinger

Julia Shawarba

Bart Robert Thomson

Andreas Wassmer

Lucas Moritz Wiggenhauser

Maria Wostrack

Sandra Wymann

Yesim Yildiz

