## Supplemental Methods for "Cerebrospinal fluid hemolysis precedes secondary brain injury after aneurysmal subarachnoid hemorrhage"

RMB, KA, SW, and SMP share first authorship; FV, MH, TG and DJS share last authorship

[1] Division of Internal Medicine, Universitätsspital and University of Zurich, Zurich, Switzerland

[2] Department of Neurosurgery, Clinical Neuroscience Center, Universitätsspital and University of Zurich, Zurich, Switzerland

[3] CSL, CSL Biologics Research Center, Bern, Switzerland

[4] Department of Biostatistics at the Epidemiology, Biostatistics and Prevention Institute, University of Zurich, Zurich, Switzerland

[5] CSL, Research & Development, Schlieren, Switzerland

### SUPPLEMENTAL METHODS

#### Targeted LC-MS/MS quantification of CSF proteins

**Overview and design.** An 8-plex LC-MS/MS MRM assay was developed for absolute quantification of eight human proteins in CSF — haptoglobin (Hp), hemopexin (Hpx), albumin, transferrin, carbonic anhydrase-1 (CAH1), bisphosphoglycerate mutase (BPGM), neurofilament light chain (NfL) and S100B. For each protein, quantification relied on one or two proteotypic surrogate peptides generated by tryptic digestion; absolute concentrations were derived from matrix-matched external calibration normalized to stable-isotope-labeled (heavy) versions of the same surrogate peptides used as internal standards. Surrogate peptides and their quantifier transitions are listed in Supplemental Table 2.

**Reference materials and internal standards.** Purified or recombinant human protein standards were obtained for all eight analytes (Hp 1-1 and Hpx, CSL Behring; albumin, CSL Behring; holo-transferrin and recombinant CAH1, RD Systems; recombinant BPGM, Abbexa; recombinant NfL, Abcam; recombinant S100B, AcroBiosystems) and stored at −70 °C. Stable-isotope-labeled surrogate peptides (Biosynth) carrying a heavy C-terminal Arg or Lys were used as internal standards (IS) for each protein; two peptides were synthesized for Hp, CAH1 and NfL. Lyophilized IS peptides were reconstituted in 50% water / 50% acetonitrile 0.1% formic acid, serially diluted, and combined into a single IS working mixture that was aliquoted and stored at −70 °C. A fixed 10-µL volume of this IS mixture was added per sample, delivering between 240 and 5000 fmol of each peptide per sample (Supplemental Table 2).

**Calibration and quality-control design.** Thirteen-point calibration curves (plus blank and double-blank) were prepared in artificial CSF (BioIVT) as a surrogate, analyte-free matrix, spanning each protein's validated range by serial dilution. Quality-control (QC) samples at low, medium and high concentrations were prepared in pooled human CSF (Innovative Research) to match the study matrix; endogenous background measured in unspiked pooled CSF was subtracted to obtain the spiked concentrations. Calibrators and QCs were prepared fresh for each analytical run. During sample acquisition, QC sets bracketed the run and were interspersed after every 10–15 study samples and after the final sample.

**Automated sample preparation.** Digestion was automated on an Agilent AssayMAP Bravo liquid handler using the "In-solution Digestion: single plate" protocol, with total hands-off processing under 2 h. Per sample, 10 µL CSF (or calibrator/blank/QC) was spiked with 10 µL IS working mixture (double-blank was spiked with 10 µL ammonium bicarbonate); proteins were then denatured by addition of 140 µL denaturation solution (50 mM ammonium bicarbonate containing sodium deoxycholate, final 0.36%) and incubation at 70 °C for 20 min. After cooling to 37 °C, 30 µL sequencing-grade modified trypsin (Promega; 0.5 µg/µL) was added and digestion proceeded at 37 °C for 60 min. Reactions were quenched with 10 µL 10% formic acid (25 °C, 10 min), which also precipitates deoxycholate. Plates were centrifuged (4,000 rpm, 15 min, room temperature) and 100 µL of supernatant transferred to a clean 96-well plate; the centrifugation/transfer step was repeated where residual deoxycholate was suspected to avoid assay interference.

**Liquid chromatography.** Peptides were separated on a Waters ACQUITY UPLC M-Class microflow system fitted with a nanoEase M/Z HSS T3 column (100 Å, 1.8 μm, 300 μm × 150 mm) held at 45 °C, at a flow rate of 10 μL/min. Mobile phase A was 0.1% formic acid in water and B was 0.1% formic acid in acetonitrile. The 17-min gradient ran 10% B (0–2 min), stepped to 15% (2.5–4.5 min), 20% (6–8 min) and 30% (8.25–9 min), ramped to 70% B at 10.5 min and 90% B at 11 min (held to 14 min), then returned to 10% B for re-equilibration to 17 min. Injection volume was 5 μL. Column effluent was diverted to waste from 0–2 min and from 10.5–17 min, with MS acquisition over the 2–10.5 min window.

**Mass spectrometry.** Detection used a SCIEX QTRAP 6500+ operated in positive-ion scheduled MRM mode (detection window 50 s) with an OptiFlow 1–50 μL Micro ion source: ion-spray voltage 4,500 V, curtain gas 30, ion source gas-1 30, low collision gas. Each protein was monitored through one or two surrogate peptides with quantifier and qualifier product-ion transitions; light (endogenous) and heavy (IS) transitions were acquired in parallel. Analyte concentrations were calculated from light/heavy peak-area ratios against the calibration curve (Supplemental Table 2).

**Analytical validation.** The assay was characterized over three independent analytical days against ICH Q2 (R1) and FDA Bioanalytical Method Validation (2018) criteria. Accuracy (recovery rate, RR), intra-assay precision (within-run CV, CVw) and inter-assay precision (between-run CV, CVb) were assessed at LLOQ, ULOQ and low/medium/high QC levels (acceptance: RR within ±15%, ±20% at LLOQ/ULOQ; CV ≤ 15%, ≤ 20% at LLOQ/ULOQ; ≥ 67% of QCs compliant). Linearity required $r^2 \geq 0.985$ with ≥ 75% and ≥ 6 non-zero calibrators meeting accuracy criteria. LLOQ required signal-to-noise (S/N) 5 and ≥ 50% of LLOQ replicates within ±20% of nominal. Specificity required blank-matrix response ≤ 20% of mean LLOQ response with S/N 5. Dilution integrity was tested at 2× and 5× dilution of above-ULOQ samples (≥ 5 replicates per factor). The matrix effect was tested in pooled human CSF spiked with 0.5%, 1%, and 2% hemolyzed whole blood at all three QC levels. On-instrument stability compared QCs injected at the start and re-injected at the end of each run. Carryover compared analyte response in water injections interspersed between samples against mean LLOQ response (acceptance ≤ 20%). All eight analytes met every predefined acceptance criterion; analyte-specific performance is summarized in Supplemental Table 3.

#### Supplemental Statistical Methods

**Analysis-set construction.** Analyses were restricted to CSF sampled via external ventricular drain. For the concurrent (same-day) GEE, the risk set comprised all clinically assessable patient-days with paired CSF measurements (one row per patient-day, including days after a patient's first SBI). For prospective first-event analyses, risk sets were horizon-specific. For H = 0, rows on the first-SBI day were retained to estimate concurrent first-event associations. For all H > 0 horizons, rows on the first-SBI day were excluded; a sampled patient-day d was eligible only if the patient was event-free through day d. Censoring then followed the rule *Y = 1 or d + H ≤ last assessed day*. A row on which first SBI occurred within the horizon window was retained irrespective of how long the patient was subsequently followed, because its outcome is already observed; only non-event rows required documented assessment through day d+H. Two further exclusions applied. Patients whose first SBI preceded their first available EVD-CSF sample were excluded (12 patients, 76 sampled patient-days, all of which fell after their first SBI), leaving 241 patients eligible at H = 0. Rows with d > 14 − H were dropped because no non-event outcome could be ascertained within the 14-day window, removing 0, 34, 75 and 123 rows at H = 0, +24, +48 and +72 hours, of which 0, 0, 1 and 4 were positives with no observable non-event counterpart. For H > 0, the horizon outcome was coded as Y_H (d) = 1 if first SBI occurred strictly after sampling, on days d+1 through d+H, and Y_H (d) = 0 if the patient remained SBI-free through d+H. Thus, no same-day first-SBI row contributed to the +24-, +48-, or +72-hour models. Rows with non-assessable SBI status were excluded from all inferential models but retained for descriptive trajectory and event-centered displays.

**Normalization and quality control.** Each biomarker was standardized independently with a skew-aware procedure. Skewness was estimated on quantile-stabilized values (1st–99th percentile, used only for the decision statistic); when the absolute skew exceeded a fixed threshold, a Yeo–Johnson power transform (which accommodates non-positive values) was fitted on the observed non-missing values before standardization. Values were then centered and scaled and clipped to ±8 standardized units to bound the influence of extreme observations, no value reached this bound (maximum |z| = 6.7). Missing values were preserved throughout (no lower-limit-of-quantification imputation), and per-biomarker quality-control plots (distribution, transform diagnostics, and clipping summaries) were generated for every analyte.

**Pathway composite scores.** For each prespecified pathway, the composite score on a given patient-day was the row-wise mean of the available standardized constituent biomarkers, computed only when at least two constituents were non-missing. The resulting mean score was re-standardized to zero mean and unit SD, so that the regression coefficient corresponds to the change in log-odds per 1-SD increment in the pathway score (OR = e^β). This re-standardization was performed once per analysis frame and was never repeated inside an individual model: the 1,925-row inferential parent set carries one scaling, which the same-day, first-event horizon and landmark models all inherit unchanged, and the 2,411-row descriptive set (which additionally contains the non-assessable patient-days) carries its own. Because the horizon and landmark risk sets are subsets of the parent set that inherit rather than recompute the score, a 1-SD increment denotes the same quantity in **Figures 2**, **3** and **4**. Both event-centered panels draw on the 2,411-row composite, but only **Figure 5B** displays it in those z units; **Figure 5A** applies a further within-patient min–max transformation to 0–1, so its cells encode each patient's position within their own range rather than a composite value. Neither panel is on the same numerical scale as the model estimates. **Figure 1F** does not use the composite at all: it plots the daily median of each constituent analyte's z-score, with the bold line the unweighted mean of those per-analyte medians. Because the analyte z-transforms were fitted once across all 2,411 EVD samples, the analyte-level z values are identical in both frames; only the composite re-standardization differs.

**Generalized estimating equations.** Logistic GEE models (binomial family, logit link) were fitted with patients as clusters and robust (sandwich) standard errors. The linear predictor was logit P (Y = 1) = α + day^ (cat) + β· (biomarker or pathway score) + site. Day was modeled with categorical dummies; when a day scheme induced perfect separation, the day factor was progressively coarsened (14 → 12 → 7 → 5 → 4 → 3 levels) and the finest separable scheme was retained. Site was entered as fixed effects, with sites contributing fewer than five patients pooled into a combined "Other" category to maintain estimability; this pooling rule was patient-based and therefore outcome-independent and constant across horizons. In this cohort no site fell below the five-patient threshold (smallest site, 15 patients), so the pooling map was empty and every site entered as its own fixed effect; the fitted models are consequently identical to a no-pooling specification by construction, and no separate refit was required. Separately from this pooling rule, day and site strata without outcome contrast were dropped before fitting at each horizon. Site E, which recorded no first SBI at any horizon, was removed from all models; day 14 was additionally removed at H = 0. Because pruning is applied after predictor-specific missingness exclusions, the number of patient-days removed depends on how completely a given predictor was measured, and is largest for the fully observed composites: 137, 102, 94 and 84 patient-days at H = 0, +24, +48 and +72 hours for hemolysis and the erythrophagocyte response, against 131, 84, 79 and 69 for neuroglial injury, whose rows are more often already absent. The retained day grid narrowed from 13 levels at H = 0 to 11 at +72 hours for four of the five composites; for neuroglial injury, day 13 also lacked contrast at H = 0, leaving 12 levels. The concurrent GEE used an exchangeable working correlation as the primary specification, with an AR (1) structure as a sensitivity analysis; the prospective horizon GEE used the same exchangeable specification; the landmark models were fitted as separate per-day logistic GLMs, with a patient-level cluster bootstrap used to account for cross-day patient overlap. Model adequacy was examined with randomized quantile residuals, calibration curves, and GEE fit summaries.

**Prospective first-event horizon construction.** At each retained patient-day, separate horizon outcomes were constructed: H = 0 captured first SBI on the sampling day, whereas the +24-, +48-, and +72-hour outcomes captured first SBI occurring strictly after the sampling day. Separate models were fitted per predictor and horizon. Benjamini–Hochberg FDR was computed across the five pathways within each horizon; H = 0 and +72-hour were prespecified as the primary horizons, and the intermediate horizons (+24 h, +48 h) were interpreted descriptively for continuity.

**Biomarker clustering.** For the secondary biomarker-level characterization, the four-horizon log-OR vector of each of the 19 analytes was clustered by Ward-linkage hierarchical clustering. The resulting ordering is a descriptive display of how the horizon profiles group, and no inferential statistic was computed on the ordering itself. Benjamini–Hochberg q values across the 19 analytes were computed within each horizon and are reported in the deposited result tables; pathway-level FDR nonetheless remained the basis for the study's inferential claims.

**Day-wise landmark analysis and cluster bootstrap.** At each landmark day $t \in \{1, \ldots, 11\}$, a logistic GLM was fitted with the day's pathway composite score as predictor and site dummies, using the risk set of patients still event-free at day t and the outcome of first SBI within (t, t + 72 h]; days with sparse counts fell back to L2-ridge penalization. Per-day log-ORs were pooled by inverse-variance weighting using the unpenalized days with finite Wald-type standard errors. Uncertainty was estimated by a patient-level cluster bootstrap (B = 2,000) that resampled whole patients with replacement (assigning pseudo-identifiers to duplicate copies), rebuilt the entire landmark cascade, and re-pooled the per-day estimates. Two quantities were derived from the resampling distribution: percentile 95% confidence intervals, and a bootstrap standard error on the pooled log-OR. Reported significance uses the Gaussian bootstrap p value, $2\Phi(-|\beta|/SE\_boot)$, Benjamini–Hochberg-adjusted across the five pathways. The two-sided percentile bootstrap p value is retained as a diagnostic only: it is bounded below by 1/B (0.0005 when all 2,000 replicates are valid, as here), and hemolysis reached that bound (p = 0.0005) while the erythrophagocyte response did not (p = 0.0010), yet Benjamini–Hochberg adjustment assigned both a common q of 0.0025, so the percentile route cannot separate the two leading pathways. A master seed (202501) deterministically seeded per-pathway and per-replicate random number generators for reproducibility. The naive independence (inverse-variance) estimate, which ignores cross-day patient overlap, is reported alongside as a reference.

**Descriptive temporal analyses.** Pathway trajectories over days 1–14 were summarized by daily medians. For the event-centered displays, patient-days were re-aligned to each patient's clinically documented first-SBI day, taken from the clinical record rather than from the sampling days, so that event timing does not depend on whether a sample happened to be drawn on the event day. The two event-centered panels are scaled differently: the per-patient heatmap (**Figure 5A**) is min–max normalized within patient to a 0–1 range across the days up to and including the event day, whereas the median trajectory (**Figure 5B**) plots the composite score in its original z units and is not normalized. Non-assessable patient-days contributed to trajectory shape and to the composite standardization, but not to event timing.

**Software and reproducibility.** Analyses used Python 3.10.10 with numpy, pandas, statsmodels (GEE), scikit-learn (ridge logistic regression), scipy, patsy, and matplotlib. Exact package versions are pinned in the requirements.txt and environment.yml files provided with the code and are logged for each run to runtime_environment.txt. A single fixed master seed (202501) makes every stochastic step reproducible: the patient-level cluster bootstrap used for the landmark pooled estimates and the scavenger residual plateaus, and the randomized quantile residuals in the model-diagnostic grids. Code and result tables are available on Zenodo (https://doi.org/10.5281/zenodo.20733246).

### SUPPLEMENTAL TABLES

**Supplemental Table 1. Sensitivity of same-day pathway associations to composite construction.** Reference is the prespecified mean composite; leave-one-marker-out (LOMO) re-estimation is shown (the most influential constituent is the marker whose removal produces the largest |ΔOR|). ORs are per 1-SD increase.

| Pathway | Dropped marker | Markers used | OR | 95% CI | p value | Reference OR | ΔOR (%) |
|---|---|---|---|---|---|---|---|
| Hemolysis | cah1_z | 3 | 1.3 | 1.05–1.60 | 0.015 | 1.31 | −1.13 |
| Hemolysis | bpgm_z | 3 | 1.31 | 1.06–1.61 | 0.013 | 1.31 | −0.64 |
| Hemolysis | hboxy_z | 3 | 1.36 | 1.09–1.69 | 0.006 | 1.31 | 3.23 |
| Hemolysis | hbmet_z | 3 | 1.3 | 1.05–1.62 | 0.018 | 1.31 | −0.96 |
| Erythrophagocytosis | cd163_z | 3 | 1.24 | 1.00–1.53 | 0.048 | 1.25 | −1.15 |
| Erythrophagocytosis | ferritin_z | 3 | 1.2 | 1.00–1.46 | 0.055 | 1.25 | −3.78 |
| Erythrophagocytosis | bilirubin_z | 3 | 1.3 | 1.06–1.61 | 0.013 | 1.25 | 4.18 |
| Erythrophagocytosis | biliverdin_z | 3 | 1.26 | 1.04–1.54 | 0.02 | 1.25 | 0.83 |
| Inflammation | crp_z | 3 | 1.29 | 1.11–1.50 | 0.001 | 1.47 | −12.44 |
| Inflammation | il6_z | 3 | 1.49 | 1.28–1.73 | <0.001 | 1.47 | 1.22 |
| Inflammation | il8_z | 3 | 1.46 | 1.24–1.72 | <0.001 | 1.47 | −0.44 |
| Inflammation | ccl2_z | 3 | 1.5 | 1.28–1.77 | <0.001 | 1.47 | 2.25 |
| Neuroglial injury | s100b_ms_z | 2 | 1.32 | 1.06–1.66 | 0.014 | 1.46 | −9.48 |
| Neuroglial injury | nse_z | 2 | 1.66 | 1.32–2.09 | <0.001 | 1.46 | 13.36 |
| Neuroglial injury | nfl_z | 2 | 1.38 | 1.13–1.70 | 0.002 | 1.46 | −5.43 |
| Plasma proteins | alb_z | 3 | 1.22 | 0.99–1.51 | 0.058 | 1.2 | 1.83 |
| Plasma proteins | trf_z | 3 | 1.21 | 0.99–1.49 | 0.066 | 1.2 | 0.95 |
| Plasma proteins | hpx_z | 3 | 1.25 | 1.01–1.54 | 0.037 | 1.2 | 3.89 |
| Plasma proteins | hp_z | 3 | 1.1 | 0.89–1.36 | 0.36 | 1.2 | −8.17 |

**Supplemental Table 2. Analytes, surrogate peptides, transitions, and calibration ranges of the HeMoVal 8-plex MRM panel.** Eight CSF proteins were quantified by MRM LC-MS/MS, each through one or two proteotypic surrogate peptides with stable-isotope-labeled internal standards. Calibrators were prepared in artificial CSF as a surrogate analyte-free matrix and quality controls in pooled human CSF with endogenous-background subtraction. The two erythrocyte enzymes (CAH1, BPGM) anchor the hemolysis composite that carries the prospective signal; the four plasma proteins constitute the comparator composite and the haptoglobin/hemopexin scavenger contrast.

| Protein | Pathway composite | Surrogate peptide(s) | Precursor charge / quantifier product ion | Heavy IS (labelled C-terminal residue) | IS spiked per sample (fmol) | Validated calibration range (LLOQ–ULOQ) |
|---|---|---|---|---|---|---|
| **Haptoglobin (HP)** | Plasma proteins | DYAEVGR (quantifier); NPANPVQ (qualifier) | 2+ / y5 (DYAEVGR) | DYAEVGR*, NPANPVQ* | 1000 (DYAEVGR); 2500 (NPANPVQ) | **0.51–340.06 µg/mL** |
| **Hemopexin (HPX)** | Plasma proteins | NFPSPVDAAFR | 2+ / y9 | NFPSPVDAAFR* | 650 | **0.72–486.77 µg/mL** |
| **Albumin (ALB)** | Plasma proteins | LVNEVTEFAK | 2+ / y6 | LVNEVTEFAK* | 5000 | **44.12–3703.70 µg/mL** |
| **Transferrin (TRF)** | Plasma proteins | GDVAFVK | 2+ / y5 | GDVAFVK* | 2000 | **0.75–506.20 µg/mL** |
| **Carbonic anhydrase-1 (CAH1)** | Hemolysis | VLDALQAIK; YSSLAEAASK | 2+ / y7 (VLDALQAIK); 2+ / y8 (YSSLAEAASK) | VLDALQAIK*, YSSLAEAASK* | 1000 (VLDALQAIK); 500 (YSSLAEAASK) | **0.18–60.01 µg/mL** |
| **Bisphosphoglycerate mutase (BPGM)** | Hemolysis | IAPEVLR | 2+ / y5 | IAPEVLR* | 1000 | **14.60–2711.21 ng/mL** |
| **Neurofilament light (NEFL/NfL)** | Neuroglial injury | ALYEQEIR (quantifier); FTVLTESAAK (qualifier) | 2+ / y6 (ALYEQEIR) | ALYEQEIR*, FTVLTESAAK* | 240 each | **29.86–554.59 ng/mL** |
| **S100B** | Neuroglial injury | AMVALIDVFHQYSGR | 3+ / y7 | AMVALIDVFHQYSGR* | 5000 | **74.08–2487.56 ng/mL** |

**Supplemental Table 3. Analytical-validation performance of the HeMoVal 8-plex MRM panel.** Assay characterization followed ICH Q2 (R1) and FDA Bioanalytical Method Validation guidance over three independent analytical days. All eight analytes met every predefined acceptance criterion; representative lower-limit performance and the validated limits are shown. Matrix effect was assessed in pooled human CSF spiked with 0.5%, 1% and 2% hemolyzed whole blood, which is the interference scenario specific to post-hemorrhagic CSF. Recovery of every quantified peptide remained within ±15%.

| Protein (quantifier peptide) | LLOQ | ULOQ | Linearity ($r^2$) | Accuracy at LLOQ (RR %) | CVb at LLOQ (%) | S/N at LLOQ | Specificity (blank/LLOQ %) | Carryover (water/LLOQ %) | Matrix effect, (RR % range) | All criteria met |
|---|---|---|---|---|---|---|---|---|---|---|
| HP (DYAEVGR) | 0.51 µg/mL | 340.06 µg/mL | ≥ 0.985 | 104.8 | 5.2 | 12.9 | 15.5 | 4.5 | 89.0–96.2 | Yes |
| HPX (NFPSPVDAAFR) | 0.72 µg/mL | 486.77 µg/mL | ≥ 0.985 | 104.4 | 3.0 | 19.3 | 10.4 | 11.1 | 96.3–104.6 | Yes |
| ALB (LVNEVTEFAK) | 44.12 µg/mL | 3703.70 µg/mL | ≥ 0.985 | 103.8 | 4.5 | 772.9 | 0.3 | 0.4 | 85.9–104.6 | Yes |
| TRF (GDVAFVK) | 0.75 µg/mL | 506.20 µg/mL | ≥ 0.985 | 98.3 | 5.2 | 227.3 | 0.9 | 0.7 | 86.6–99.1 | Yes |
| CAH1 (YSSLAEAASK) | 0.18 µg/mL | 60.01 µg/mL | ≥ 0.985 | 102.5–107.2 | 3.5–5.3 | 83–127 | 1.6–2.4 | 0.55–2.1 | 93.2–106.2 | Yes |
| BPGM (IAPEVLR) | 14.60 ng/mL | 2711.21 ng/mL | ≥ 0.985 | 93.0–104.6 | 9.1–10.3 | 55.2 | 3.6 | 0.0 | 82.6–99.1 | Yes |
| NfL (ALYEQEIR) | 29.86 ng/mL | 554.59 ng/mL | ≥ 0.985 | 99.0 | 9.2 | 75.8 | 2.6 | 1.3 | 91.0–114.0 | Yes |
| S100B (AMVALIDVFHQYSGR) | 74.08 ng/mL | 2487.56 ng/mL | ≥ 0.985 | 102.5 | 13.5 | 19.1 | 10.5 | 0.0 | 87.3–99.8 | Yes |

.

**Supplemental Table 4. Analysis-set and sampling characteristics of the HeMoVal EVD-CSF biomarker cohort.** Full baseline demographic and severity characteristics of the enrolled cohort are reported in the registered primary analysis (25); this table summarizes the analysis-set and serial-sampling characteristics specific to the present biomarker study (derived from the deposited dataset).

| Characteristic | Value |
|---|---|
| Participating centers | 8 |
| Patients with EVD-sampled CSF | 259 |
| - with SBI | 126 (48.6%) |
| - without SBI | 127 (49.0%) |
| - non-assessable throughout | 6 (2.3%) |
| Patients contributing to inferential models | 253 |
| EVD-CSF samples | 2,411 |
| - with assessable SBI status | 1,925 (296 SBI event-days; 1,629 non-event-days) |
| - with non-assessable SBI status (descriptive only) | 486 |
| Sampling window | days 1–14 post-SAH |
| Samples per patient, all | median 10 (IQR 7–13) |
| - patients with SBI | median 12 (IQR 9–13) |
| - patients without SBI | median 9 (IQR 5–11) |

**Supplemental Table 5. Site-level distribution of patients, SBI classification, and EVD-CSF samples in the HeMoVal biomarker cohort.** The table shows the number of patients according to SBI classification and the number of EVD-CSF samples contributed by each participating center. Percentages use the 259-patient EVD denominator; the inferential set (253 patients) excludes the 6 non-assessable-throughout patients. Site labels are de-identified and sites are ordered by patient count. SBI, secondary brain injury; EVD, external ventricular drain; CSF, cerebrospinal fluid.

| Site | Patients | SBI | No-SBI | Non-assessable | CSF samples |
|---|---|---|---|---|---|
| **A** | 66 | 40 | 25 | 1 | 649 |
| **F** | 50 | 29 | 21 | 0 | 522 |
| **D** | 31 | 14 | 14 | 3 | 268 |
| **H** | 30 | 10 | 20 | 0 | 253 |
| **C** | 28 | 15 | 12 | 1 | 208 |
| **E** | 20 | 1 | 19 | 0 | 164 |
| **B** | 19 | 8 | 10 | 1 | 163 |
| **G** | 15 | 9 | 6 | 0 | 184 |

### SUPPLEMENTAL FIGURES

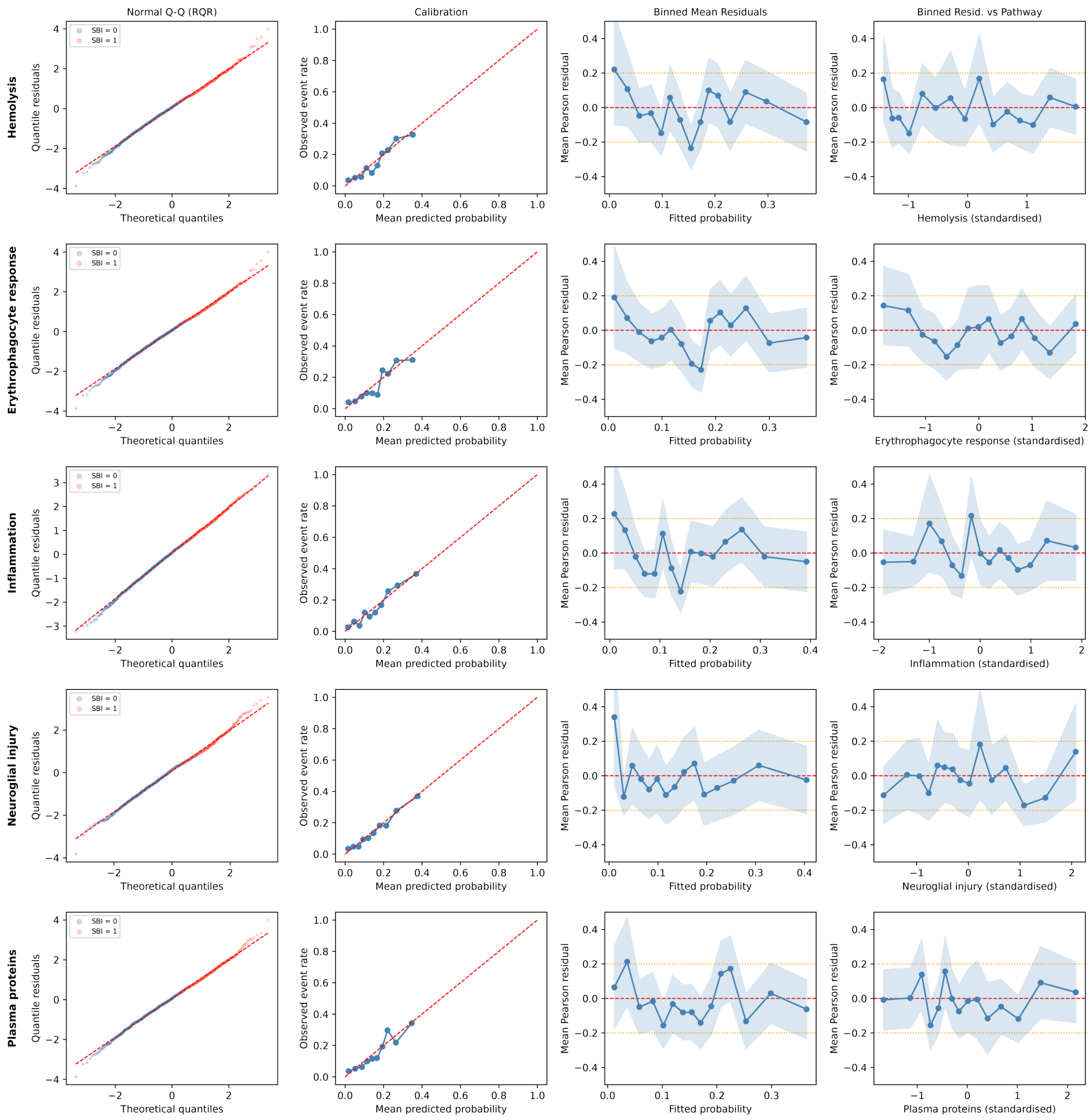


**Supplemental Figure 1. Model-diagnostic grid for the concurrent same-day pathway GEE.** Rows are the five prespecified pathway composites. Columns show, from left to right, a Normal Q–Q plot of randomized quantile residuals, a calibration curve (observed event rate versus mean predicted probability, 10 quantile bins), binned mean Pearson residuals versus fitted probability, and binned mean Pearson residuals versus the pathway score. All panels are from the primary exchangeable working-correlation specification underlying **Figure 2A**. Calibration and randomized quantile residual diagnostics were consistent with adequate fit for all five pathways. Equivalent diagnostic grids for the prospective first-event models at H = 0 and +72 hours are provided with the deposited results; these showed a mild calibration attenuation shared across all five pathways at +72 hours that did not alter the relative pathway ranking.

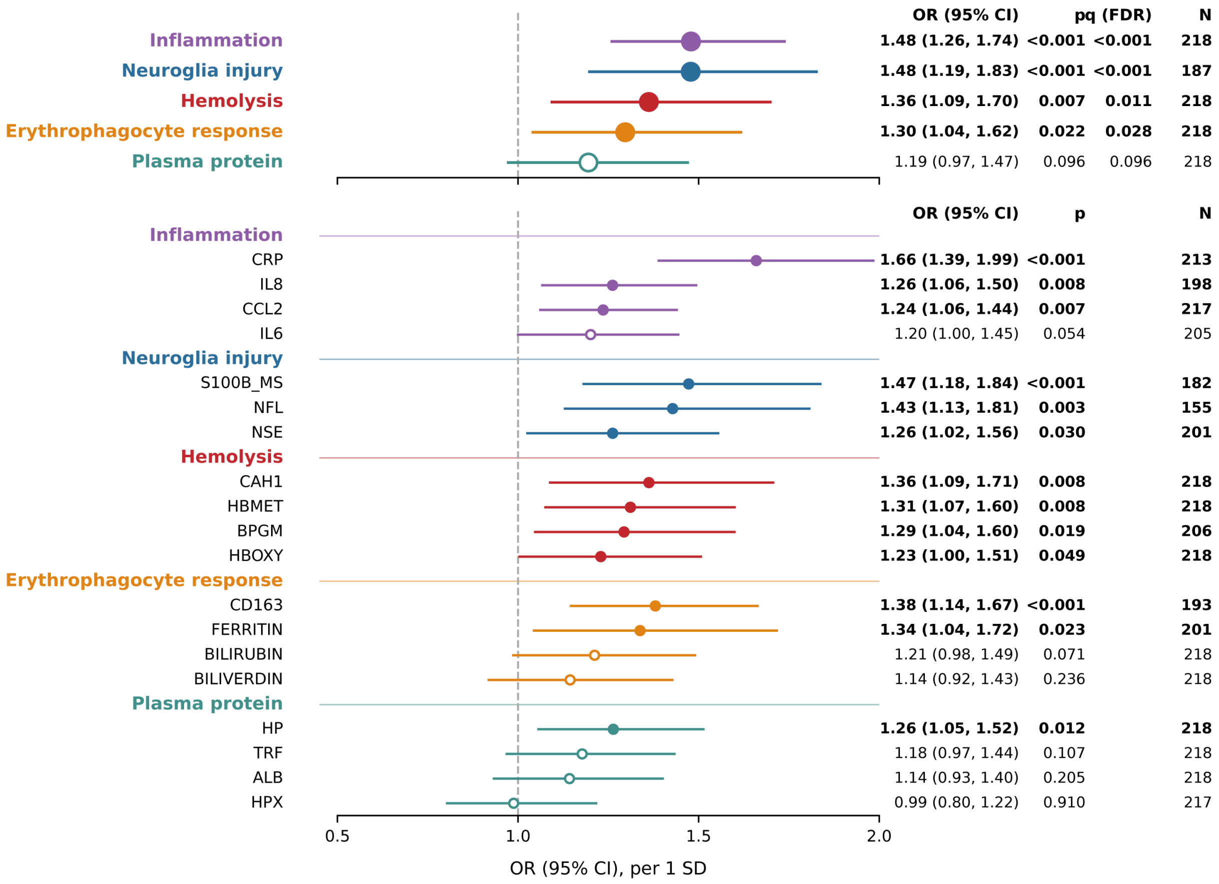


**Supplemental Figure 2. AR (1) working-correlation sensitivity for same-day associations.** Forest plot of same-day GEE odds ratios (per 1-SD increase, 95% CI) estimated under an AR (1) within-patient working correlation, shown at the pathway level and for the 19 constituent biomarkers grouped by pathway. Estimates were concordant with the primary exchangeable-correlation specification (Figure 2).

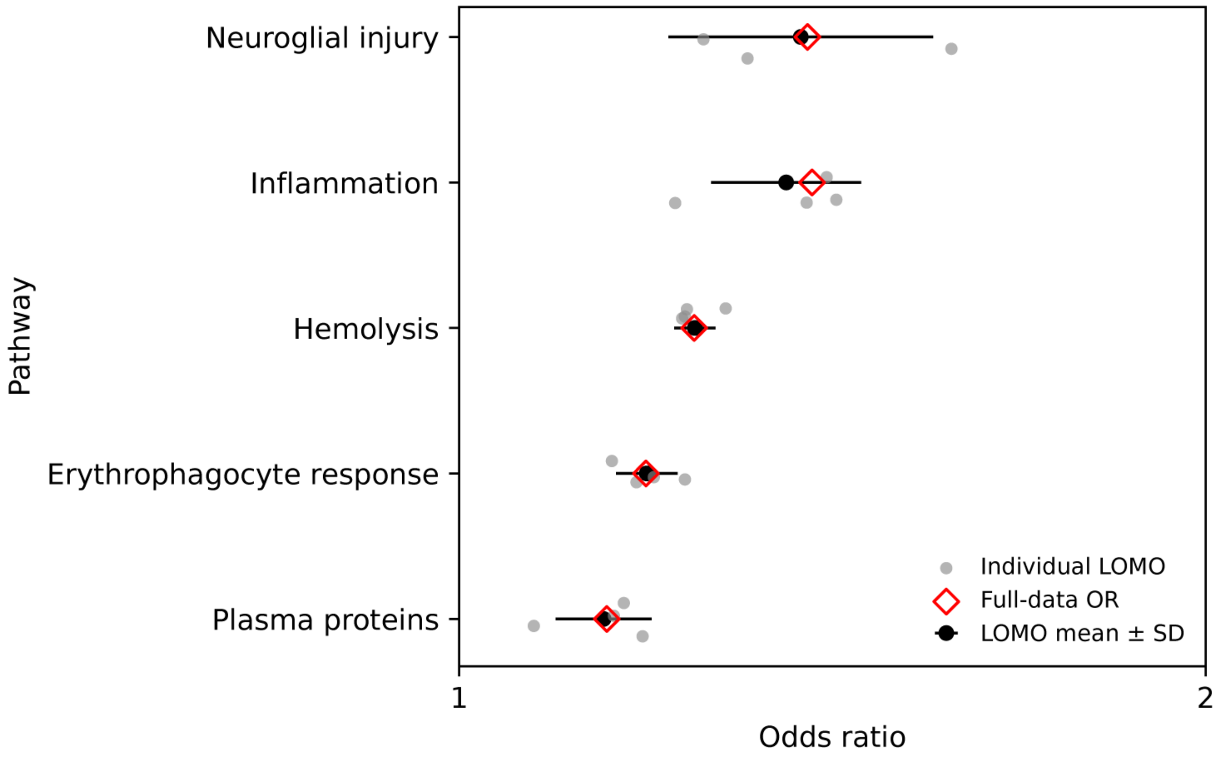


**Supplemental Figure 3. Leave-one-marker-out sensitivity analysis of same-day pathway associations.** For each pathway, grey points show odds ratios from individual leave-one-marker-out (LOMO) models, red diamonds show the full-data pathway odds ratio, and black points with horizontal bars show the mean ± SD of the LOMO estimates. The direction of association was preserved in every LOMO model for every pathway, so the spread of the leave-one-marker-out estimates reflects each constituent's contribution to effect magnitude rather than dependence on any single marker.
