## Supplementary material for "Cerebrospinal fluid hemolysis precedes secondary brain injury after aneurysmal subarachnoid hemorrhage": STROBE checklist

STROBE Statement—Checklist of items that should be included in reports of ***cohort studies***

| | Item No | Recommendation | Page No |
|---|---|---|---|
| **Title and abstract** | 1 | (*a*) Indicate the study's design with a commonly used term in the title or the abstract | 2 |
| | | (*b*) Provide in the abstract an informative and balanced summary of what was done and what was found | 2 |
| **Introduction** | | | |
| Background/rationale | 2 | Explain the scientific background and rationale for the investigation being reported | 3-4 |
| Objectives | 3 | State specific objectives, including any prespecified hypotheses | 3-4; 11 |
| **Methods** | | | |
| Study design | 4 | Present key elements of study design early in the paper | 2-4; 11-12 |
| Setting | 5 | Describe the setting, locations, and relevant dates, including periods of recruitment, exposure, follow-up, and data collection | 11-12; recruitment dates not reported |
| Participants | 6 | (*a*) Give the eligibility criteria, and the sources and methods of selection of participants. Describe methods of follow-up | 11-12; 20-21 (Fig. 1); Suppl. pp. 3-5 |
| | | (*b*) For matched studies, give matching criteria and number of exposed and unexposed | N/A - not matched |
| Variables | 7 | Clearly define all outcomes, exposures, predictors, potential confounders, and effect modifiers. Give diagnostic criteria, if applicable | 3-4; 11-14; 20-26; diagnostic criteria: protocol (ref. 24) |
| Data sources/ measurement | 8* | For each variable of interest, give sources of data and details of methods of assessment (measurement). Describe comparability of assessment methods if there is more than one group | 4; 11-13; 20-21; Suppl. pp. 2-4, 8-9 |
| Bias | 9 | Describe any efforts to address potential sources of bias | 4; 7; 10-15; Suppl. pp. 3-6, 11 |
| Study size | 10 | Explain how the study size was arrived at | 12; Suppl. p. 10 (available cohort; size rationale partial) |
| Quantitative variables | 11 | Explain how quantitative variables were handled in the analyses. If applicable, describe which groupings were chosen and why | 13-15; Suppl. pp. 4-6 |

| | | | |
|---|---|---|---|
| Statistical methods | 12 | (*a*) Describe all statistical methods, including those used to control for confounding | 5-7; 13-15; 22-24; Suppl. pp. 3-6 |
| | | (*b*) Describe any methods used to examine subgroups and interactions | No subgroup or interaction analyses reported |
| | | (*c*) Explain how missing data were addressed | 4; 11-14; Suppl. pp. 3-5 |
| | | (*d*) If applicable, explain how loss to follow-up was addressed | 10-11; 14; Suppl. pp. 3-4 |
| | | (*e*) Describe any sensitivity analyses | 5; 7; 14-15; Suppl. pp. 7, 12-13 |
| **Results** | | | |
| Participants | 13* | (a) Report numbers of individuals at each stage of study—eg numbers potentially eligible, examined for eligibility, confirmed eligible, included in the study, completing follow-up, and analysed | 4; 12; 20-21; Suppl. pp. 3-5, 10 (partial flow) |
| | | (b) Give reasons for non-participation at each stage | 12; 20-21; Suppl. pp. 3-5 (partial reporting) |
| | | (c) Consider use of a flow diagram | 20-21 (Fig. 1) |
| Descriptive data | 14* | (a) Give characteristics of study participants (eg demographic, clinical, social) and information on exposures and potential confounders | 11; 15-16; Suppl. p. 10 (partial; baseline data: ref. 25) |
| | | (b) Indicate number of participants with missing data for each variable of interest | 4; 13; 20-21; Suppl. p. 10 (sample-level only) |
| | | (c) Summarise follow-up time (eg, average and total amount) | 11-12; 14; Suppl. p. 10 (window only; duration not reported) |
| Outcome data | 15* | Report numbers of outcome events or summary measures over time | 4-8; 10; 20-21; 26; Suppl. p. 10 |

| | | | |
|---|---|---|---|
| Main results | 16 | (*a*) Give unadjusted estimates and, if applicable, confounder-adjusted estimates and their precision (eg, 95% confidence interval). Make clear which confounders were adjusted for and why they were included | 5-8; 22-26; Suppl. p. 7 (adjusted only; unadjusted not reported) |
| | | (*b*) Report category boundaries when continuous variables were categorized | 13-14; Suppl. pp. 4-5 (continuous biomarkers; categorical day) |
| | | (*c*) If relevant, consider translating estimates of relative risk into absolute risk for a meaningful time period | Not reported |
| Other analyses | 17 | Report other analyses done—eg analyses of subgroups and interactions, and sensitivity analyses | 5-8; 14-15; 23-26; Suppl. pp. 7, 12-13 |
| **Discussion** | | | |
| Key results | 18 | Summarise key results with reference to study objectives | 8; 11 |
| Limitations | 19 | Discuss limitations of the study, taking into account sources of potential bias or imprecision. Discuss both direction and magnitude of any potential bias | 10-11 |
| Interpretation | 20 | Give a cautious overall interpretation of results considering objectives, limitations, multiplicity of analyses, results from similar studies, and other relevant evidence | 8-11 |
| Generalisability | 21 | Discuss the generalisability (external validity) of the study results | 10 |
| **Other information** | | | |
| Funding | 22 | Give the source of funding and the role of the funders for the present study and, if applicable, for the original study on which the present article is based | 2; 16 |
